# Liver disease is a significant risk factor for cardiovascular outcomes - a UK Biobank study

**DOI:** 10.1101/2022.12.08.22283242

**Authors:** Adriana Roca-Fernandez, Rajarshi Banerjee, Helena Thomaides-Brears, Alison Telford, Arun Sanyal, Stefan Neubauer, Thomas E Nichols, Betty Raman, Celeste McCracken, Steffen E Petersen, Ntobeko AB Ntusi, Daniel J Cuthbertson, Michele Lai, Andrea Dennis, Amitava Banerjee

## Abstract

**Background:** Chronic liver disease (CLD) and cardiovascular diseases (CVD) share common risk factors; the former is associated with a two-fold greater incidence of CVD. With most CLD being preventable/modifiable, early identification of at high-risk individuals is crucial. Using data from the UK Biobank imaging sub-study, we tested the hypothesis that early signs of liver disease (measured by iron corrected T1-mapping (cT1)) is associated with an increased risk of major cardiovascular events.

**Methods:** Liver disease activity (cT1) and fat (PDFF) were measured using LiverMultiScan® from images acquired between January-2016 and February-2020 in the UK Biobank imaging sub-study. Multivariable Cox regression was used to explore associations between liver cT1 (MRI) and ***primary CVD outcomes*** (coronary artery disease, atrial fibrillation, embolism/vascular events, heart failure and stroke), as well as CVD ***hospitalisation*** and ***all-cause mortality***. Other liver blood biomarkers (AST, ALT, AST/ALT ratio, FIB4), general metabolism biomarkers (CRP, HbA1c, systolic blood pressure (SBP), total cholesterol), and demographics were also included. Subgroup analysis was conducted in those without metabolic syndrome (MetS= at least 3 of these traits: a large waist, high triglycerides, low HDL cholesterol, increased SBP, or elevated HbA1c)

**Results:** 33,616 participants in the UK Biobank imaging sub-study (65 years, mean BMI 26kg/m^2^, mean HbA1c 35mmol/mol) had complete MRI liver data with linked clinical outcomes [median time to major CVD event onset: 1.4 years (range:0.002-5.1); follow-up: 2.5 years (range:1.1-5.2)]. Liver disease activity (cT1), but not liver fat (PDFF), was associated with a higher risk of any major CVD event [HR(CI) 1.14(1.03-1.26), p=0.008], AF [1.30 (1.12-1.5), p<0.001]; HF [1.30 (1.08 - 1.58), p=0.004]; CVD hospitalisation [1.27(1.18-1.387, p<0.001] and all-cause mortality [1.19(1.02-1.38), p=0.026]. FIB4 index, was associated with HF [1.06 (1.01 - 1.10)), p=0.007]. The risk of CVD hospitalisation was also independently associated with cT1 in individuals without MetS [1.26(1.13-1.4), p<0.001].

**Conclusion:** Liver disease activity, as measured with MRI-derived biomarker cT1, was independently associated with a higher risk of new onset CVD events and all-cause mortality. This association occurred even without pre-existing impairment of metabolic health and was independent of FIB4 or liver fat content. cT1 was identified as a major predictor of adverse CVD outcomes.

## INTRODUCTION

Over the past decade, incidence of chronic liver disease (CLD) related to non-alcoholic metabolic-associated fatty liver disease (NAFLD/MAFLD) has been increasing. NAFLD, affecting 25% of the population globally, is now the principal driver of cirrhosis, hepatocellular cancer and liver transplantation (1,2). Multiple non-invasive biomarkers for both early and late stage CLD have been associated with liver-related outcomes (3,4) and adopted in drug efficacy trials (5–8) and clinical practice (9–11). However, cardiovascular disease (CVD) is a leading cause of death in NAFLD patients (12,13); and coronary artery disease (CAD), arrhythmia, and stroke are considerably more common in most CLDs (14). As such, society guidelines are recommending CVD screening in patients with CLD (10,15,16). However specific risk-scoring based treatment algorithms are lacking in this population (10). This may be because the mechanisms behind increased CVD risk in those with NAFLD remain unclear, with inflammatory (17), metabolic and immune-mediated processes all being suggested. A recent large-scale study of electronic health records from over 4 million adults reported a consistent pattern of increased body mass index (BMI _≥_30), and comorbidities such as type 2 diabetes, hypertension, and chronic kidney disease, all associated with CVD risk across a range of liver diseases (NAFLD, alcoholic liver disease, viral and autoimmune hepatitis), as were serum/plasma based markers of inflammation such as C-reactive protein. However, in that study, abnormalities in conventional liver biochemistry (e.g. bilirubin, aspartate transaminase (AST) and gamma-glutamyl transferase (GGT)), were not associated with CVD risk (14). Various biochemistry-based risk scores have been incorporated into clinical assessment, such as AST/ALT ratio, (commonly used to differentiate causes of liver damage), the fibrosis-4 index (FIB4) (for initial screening for liver fibrosis) (9,10,18,19) and NAFLD fibrosis score; all these biomarkers predict liver-related outcomes (such as cirrhosis, liver transplant and hepatocellular carcinoma (HCC)). (20). While these biomarkers have shown to be independent predictors (21), or associated (22) with major adverse cardiovascular events, these have been tested only in populations with established diagnoses of liver disease. Thus, risk stratification for early asymptomatic liver disease and CVD clinical outcomes still needs to be further investigated (23).

Quantitative biomarkers derived from medical imaging have gathered momentum in both the cardiac and the CLD space, as they are non-invasive, allow for assessment of the whole organ and are inherently organ related. MRI-derived T1-mapping of the heart has been shown to be associated with a variety of cardiomyopathies (including diffuse fibrosis (24) and myocarditis (25)), resulting in inclusion in clinical guidelines, such as the Society for cardiovascular magnetic resonance (26). In the liver, iron-corrected T1 mapping (cT1) is a marker of CLD activity, which has been shown to correlate with parenchymal ballooning, inflammation and fibrosis (27) and has been associated with histological disease activity in steatohepatitis (28), and viral (29) and autoimmune hepatitis (30). cT1 has been shown to predict liver-related outcomes in patients with CLD (4). cT1 has since been recognised by gastroenterology and endocrinology guidelines in the assessment of patients with NAFLD (10,31). Proton density fat fraction (PDFF) is a biomarker of liver fat that can stratify all grades of liver steatosis and is used clinically for screening and as a clinical trial endpoint (7,9,32), but has not been reported to be associated with clinical events.

The UK Biobank is a large-scale biomedical database in the United Kingdom investigating the development of disease, exploring both genetic predisposition, and environmental exposure (33). We sought to explore associations between the liver and cardiovascular clinical outcomes using this resource. We investigated: 1) associations between established non-invasive (blood and imaging) CLD biomarkers and CVD outcomes, 2) how these associations related to CLD characteristics and 3) whether associations with CVD events were independent from associated metabolic disease.

## METHODS

The UK Biobank imaging sub study is an ongoing study aiming to scan the brains, hearts, bones and abdomens of 100,000 of the 502,506 UK Biobank participants (34). A retrospective analysis of all the data available as acquired between January 2016 and February 2020 was performed. UK Biobank has approval from the Northwest Multi-Centre Research Ethics Committee (MREC) and obtained written informed consent from all participants prior to the study. The data were extracted under access application 9914. Some of the authors (AR-F, RB, CM, and AD) had access to all the data through this application and take responsibility for the contents of this manuscript. Patients and public were involved at every stage of the conception and design of UK Biobank and patients with CLD contributed to this article and the patient impact of this research.

### Study Population

The inclusion criteria included those who had a complete set of liver image-derived phenotypes for liver fat (PDFF, %) and disease activity (cT1, milliseconds) from the abdominal imaging protocol. The CONSORT diagram is shown in **Figure 1**. There were no exclusion criteria. Patient meta-data including information on demographics at the time of the scan were available. Cardiometabolic risk factors and metabolic blood biomarkers associated with CLD were collected at baseline assessment.

**Figure 1:**
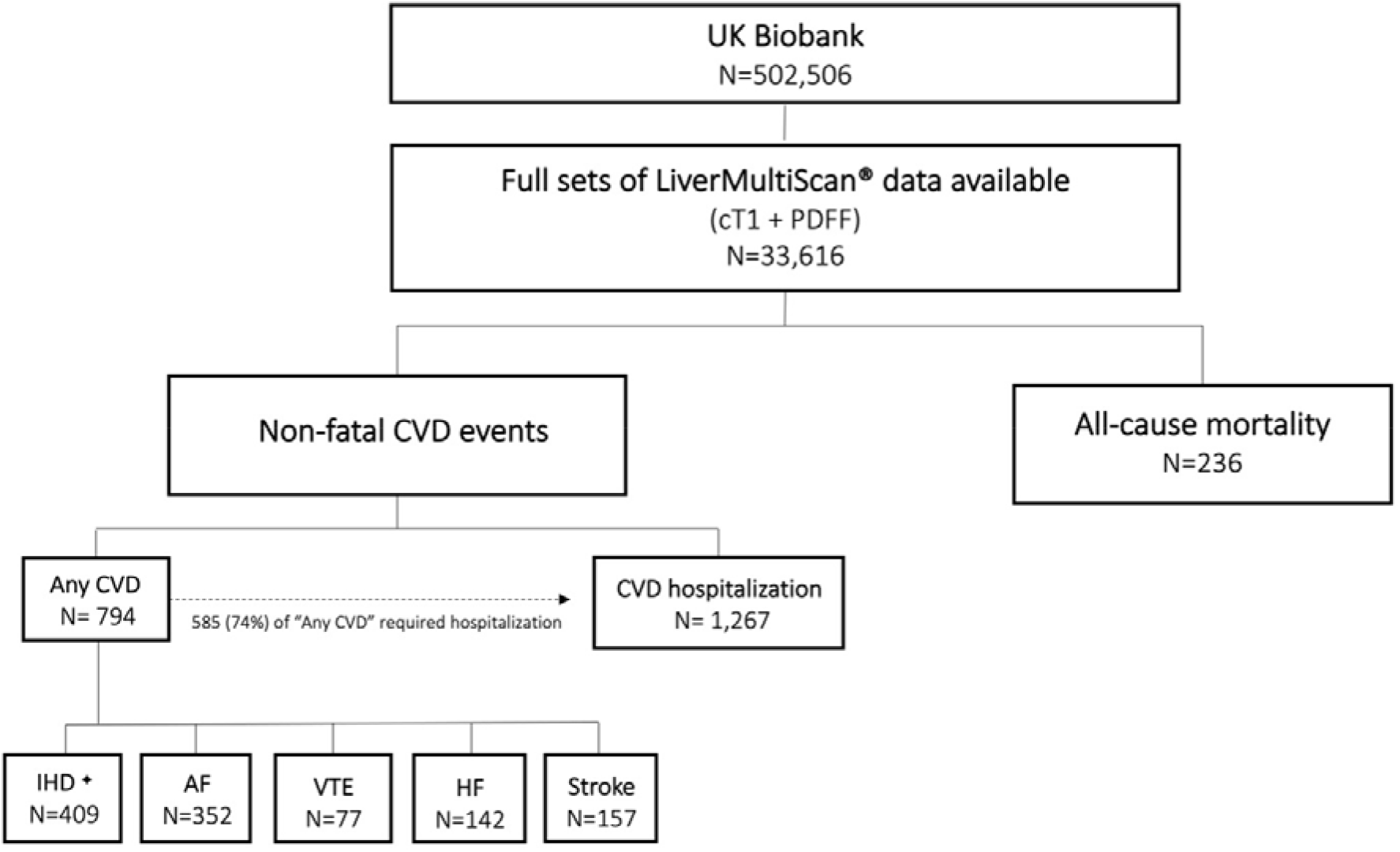
Consort diagram. Abbreviations: CVD, cardiovascular disease; CAD, coronary artery disease, ⍰ includes 160 cases of acute myocardial infarction; AF, atrial fibrillation; VTE, Embolism/vascular event; NIC, non-ischemic cardiomyopathy; HF, heart failure; (see supplementary tables 1 and 2 for more details).

### Independent variables and outcomes

New onset CVD clinical events were the outcomes of interest, specifically coronary artery disease (CAD), atrial fibrillation (AF), embolism/vascular events (VTE), heart failure (HF), and stroke. ICD-10 codes to define events were agreed by consensus cardiologists (BR, SN, RB) based on Bosco et al (35). Hospitalisation due to a primary cardiovascular event and all-cause mortality were also recorded. To ensure capture of the most severe events, these were defined as the first event for each patient following their UK Biobank imaging visit **(Supplementary table S1)**. Inpatients were defined as individuals who were admitted to hospital and occupy a hospital bed. This included both admissions where an overnight stay was planned and day cases.

Liver measurements derived from the LiverMultiScan® software and standard liver function tests (AST/ALT ratio, ALT, AST, and FIB4 index [(Age*AST)/ (Platelet count*sqrt (ALT))]) were assessed as predictors of CVD events. Additional blood biomarkers (high-density lipoprotein (HDL), low-density lipoprotein (LDL), haemoglobin A1c (HbA1C), triglycerides, total cholesterol, and C-reactive protein) were also evaluated. Previously reported risk factors which included being male, age, BMI, systolic blood pressure and smoking were also explored (36,37).

### Imaging protocol and analysis

Participants were scanned at one of four UK Biobank imaging centres using the LiverMultiScan® protocol from Perspectum Ltd (UK) which forms part of the UK Biobank abdominal imaging protocol. Liver MRI data was analysed automatically using the LiverMultiScan® software, and every case was manually reviewed by trained analysts, blinded to any subject variables. Example cT1 images are displayed in **figure 3**.

### Statistical Analysis

Statistical analysis was performed using R software (version 4.0.4) with a p-value <0.05 considered statistically significant. Descriptive statistics were used to summarize baseline participant characteristics. Mean and standard deviation (SD) were used to describe normally distributed-continuous variables, median with interquartile range (IQR) for non-normally distributed, and frequency and percentage for categorical variables. For group-wise comparisons of continuous parametric and non-parametric, and categorical variables, T-test, Wilcoxon rank sum and Fisher’s exact tests, respectively, were used.

Analyses into the associations between CDV events (CAD, AF, VTE, HF, stroke, hospitalisation, and all-cause mortality) and image biomarkers, blood biomarkers, comorbidities, and demographics was performed using Cox proportional hazard regression analysis. This was initially done univariately (each variable separately) to study the contribution of an individual variable on the occurrence of each specific clinical event. Significant variables were included in a multivariable Cox proportional hazard regression model to assess which variables are independent predictors of CVD events. This process was performed under three different conditions: (1) with all biomarkers treated as continuous variables following transformation into Z-scores, (2) using pre-defined clinically used thresholds, and (3) in individuals without comorbid metabolic syndrome (MetS). A cT1 threshold was _≥_800ms is considered the upper limit of normal and the recommended threshold to identify those in transition from simple steatosis to NASH (28). MetS was defined as having three or more of the following traits: a large waist (_≥_ 89cm waist circumference in women and 102cm in men), high triglycerides (TGA) (_≥_ 1.7mmol/L), low HDL cholesterol (<1.04mmol/L in men and < 1.3mmol/L in women), increased systolic blood pressure (_≥_ 130/85mmHg) or elevated HbA1c (_≥_ 32mmol/mol) (38).

## RESULTS

Data from 41,994 participants were extracted from the UK Biobank imaging showcase. Of these, 33,616 had full liver measurements of imaging and biochemistry. In the time following the MRI scan, 794 participants (2.4%) experienced a form of cardiovascular event requiring hospitalisation. Looking at the specific CVD categories, 409 participants experienced CAD events, (including 160 acute myocardial infarctions), 352 AF events, 77 VTE events, 142 HF events and 157 stroke events. In addition, 1267 individuals required hospitalisation due to any cardiovascular event and 236 individuals died **(Figure 1)**.

Median time to event was 1.4 years (0.002-5.1) and the median follow up time for the population was 2.5 years (1.1-5.2). Comparing those with and without any major CVD event after the MRI scan, participants experiencing events were older (p<.001), had higher BMI (p<.001) and were more likely to be male (64%, p<0.001). Participant characteristics for the whole population and relevant subgroups are reported in **Table 1**.

**Table 1:**
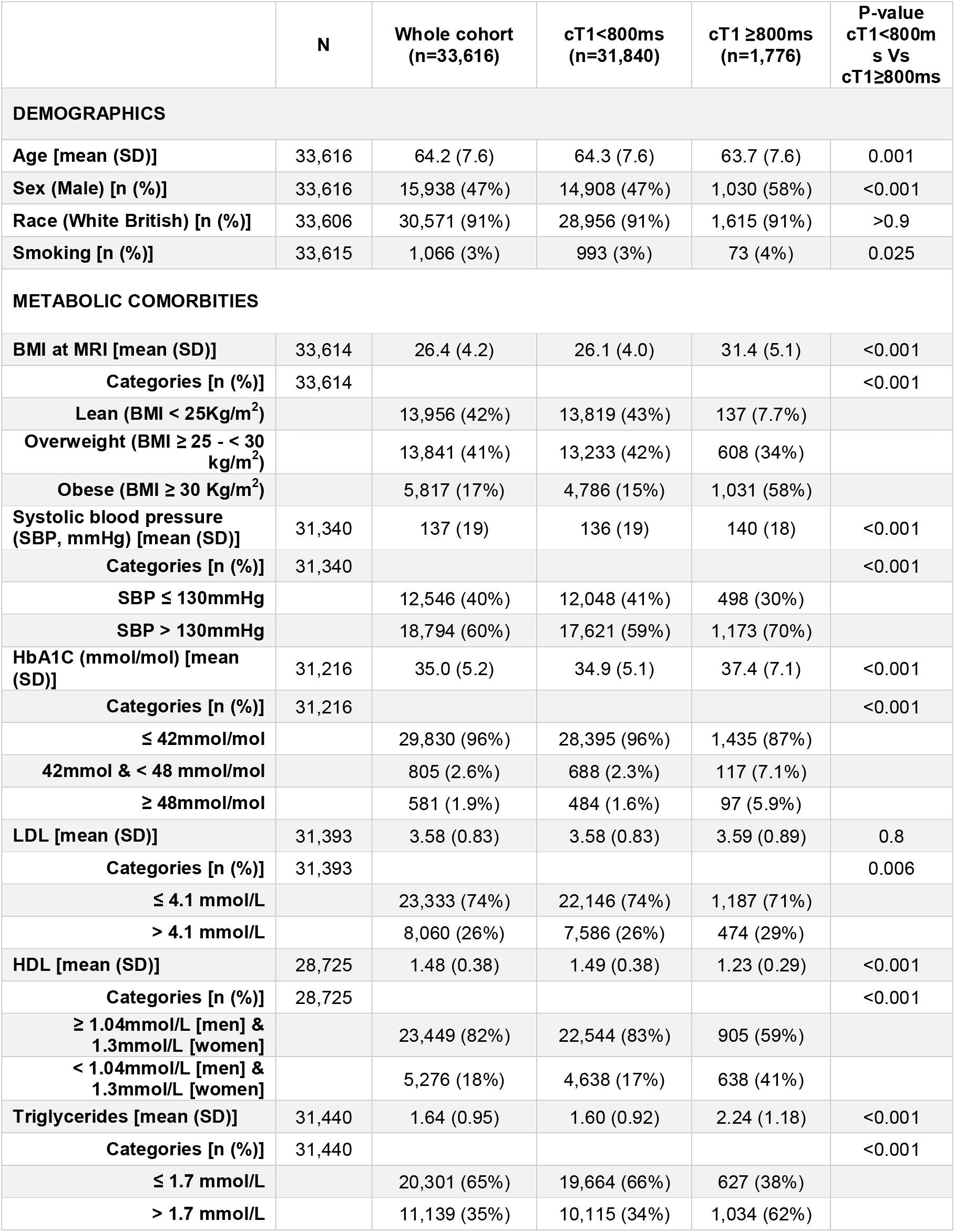
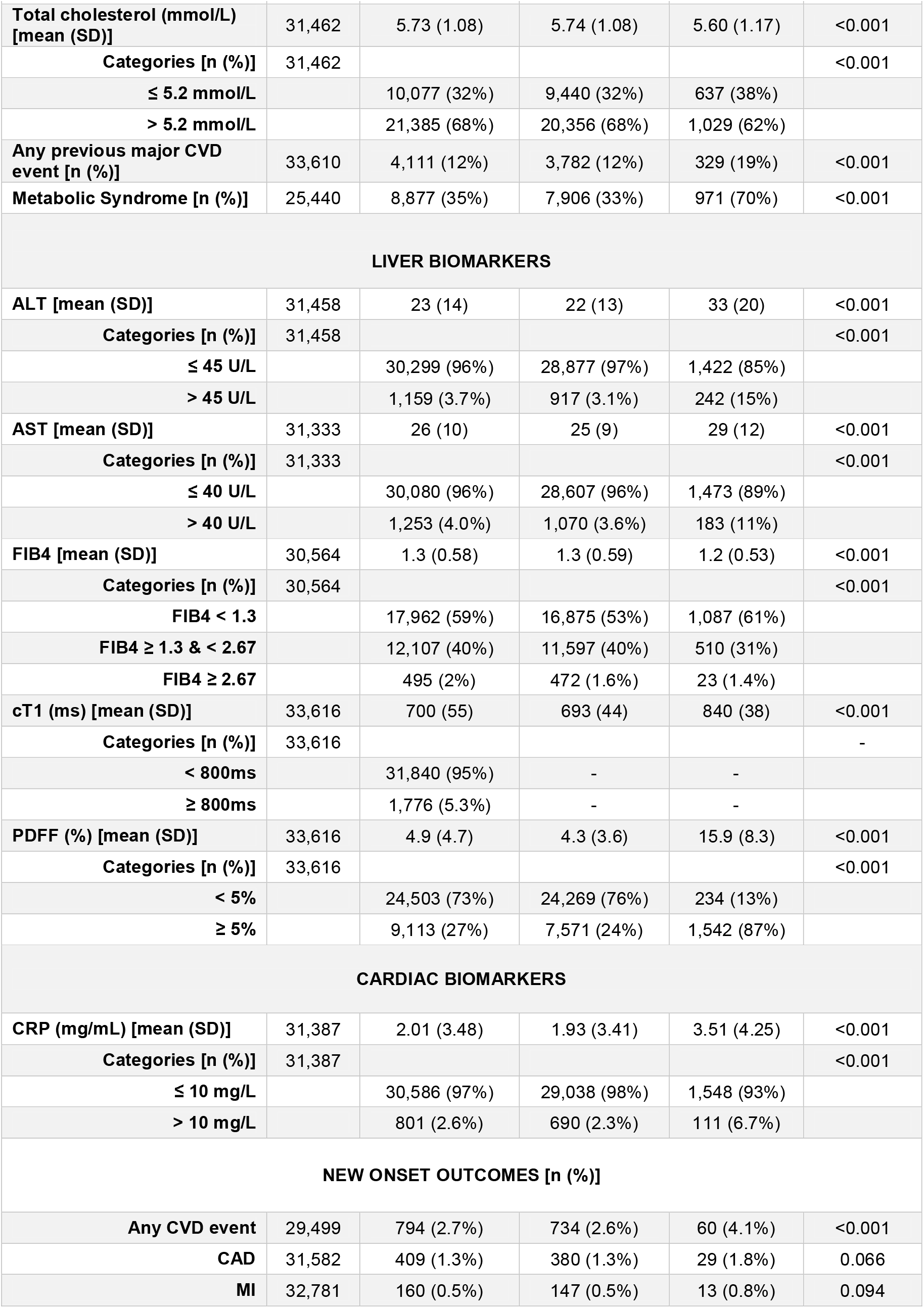
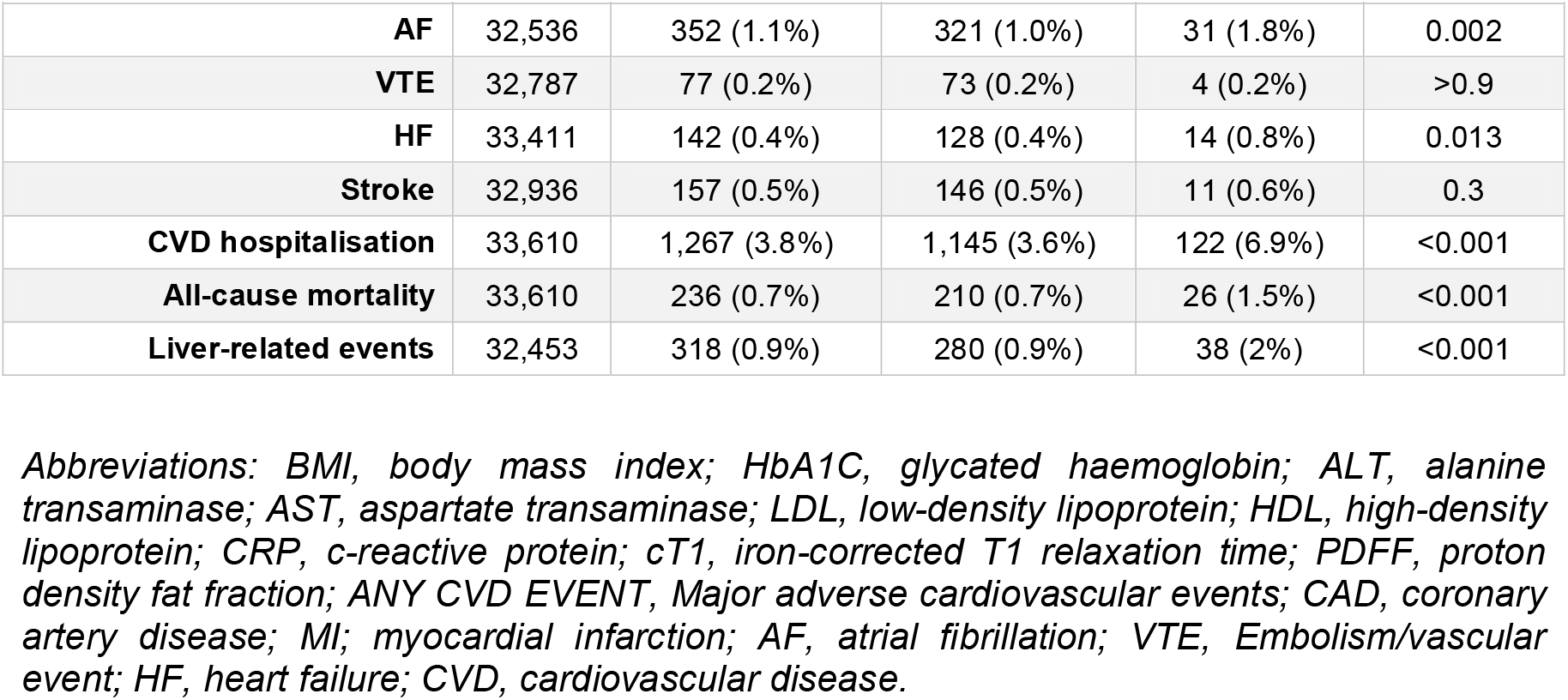
Demographics, baseline characteristics and clinical outcomes in the whole cohort and according to liver cT1. Age was at MRI visit; bloods were taken at baseline visit.

|  | N | Whole cohort<br>(n=33,616) | cT1<800ms<br>(n=31,840) | cT1 ≥800ms<br>(n=1,776) | P-value<br>cT1<800ms Vs<br>cT1≥800ms |
| --- | --- | --- | --- | --- | --- |
| **DEMOGRAPHICS** |  |  |  |  |  |
| Age [mean (SD)] | 33,616 | 64.2 (7.6) | 64.3 (7.6) | 63.7 (7.6) | 0.001 |
| Sex (Male) [n (%)] | 33,616 | 15,938 (47%) | 14,908 (47%) | 1,030 (58%) | <0.001 |
| Race (White British) [n (%)] | 33,606 | 30,571 (91%) | 28,956 (91%) | 1,615 (91%) | >0.9 |
| Smoking [n (%)] | 33,615 | 1,066 (3%) | 993 (3%) | 73 (4%) | 0.025 |
| **METABOLIC COMORBIDITIES** |  |  |  |  |  |
| BMI at MRI [mean (SD)] | 33,614 | 26.4 (4.2) | 26.1 (4.0) | 31.4 (5.1) | <0.001 |
| Categories [n (%)] | 33,614 |  |  |  | <0.001 |
| Lean (BMI < 25Kg/m<sup>2</sup>) |  | 13,956 (42%) | 13,819 (43%) | 137 (7.7%) |  |
| Overweight (BMI ≥ 25 - < 30 kg/m<sup>2</sup>) |  | 13,841 (41%) | 13,233 (42%) | 608 (34%) |  |
| Obese (BMI ≥ 30 Kg/m<sup>2</sup>) |  | 5,817 (17%) | 4,786 (15%) | 1,031 (58%) |  |
| Systolic blood pressure (SBP, mmHg) [mean (SD)] | 31,340 | 137 (19) | 136 (19) | 140 (18) | <0.001 |
| Categories [n (%)] | 31,340 |  |  |  | <0.001 |
| SBP ≤ 130mmHg |  | 12,546 (40%) | 12,048 (41%) | 498 (30%) |  |
| SBP > 130mmHg |  | 18,794 (60%) | 17,621 (59%) | 1,173 (70%) |  |
| HbA1C (mmol/mol) [mean (SD)] | 31,216 | 35.0 (5.2) | 34.9 (5.1) | 37.4 (7.1) | <0.001 |
| Categories [n (%)] | 31,216 |  |  |  | <0.001 |
| ≤ 42mmol/mol |  | 29,830 (96%) | 28,395 (96%) | 1,435 (87%) |  |
| 42mmol & < 48 mmol/mol |  | 805 (2.6%) | 688 (2.3%) | 117 (7.1%) |  |
| ≥ 48mmol/mol |  | 581 (1.9%) | 484 (1.6%) | 97 (5.9%) |  |
| LDL [mean (SD)] | 31,393 | 3.58 (0.83) | 3.58 (0.83) | 3.59 (0.89) | 0.8 |
| Categories [n (%)] | 31,393 |  |  |  | 0.006 |
| ≤ 4.1 mmol/L |  | 23,333 (74%) | 22,146 (74%) | 1,187 (71%) |  |
| > 4.1 mmol/L |  | 8,060 (26%) | 7,586 (26%) | 474 (29%) |  |
| HDL [mean (SD)] | 28,725 | 1.48 (0.38) | 1.49 (0.38) | 1.23 (0.29) | <0.001 |
| Categories [n (%)] | 28,725 |  |  |  | <0.001 |
| ≥ 1.04mmol/L [men] & 1.3mmol/L [women] |  | 23,449 (82%) | 22,544 (83%) | 905 (59%) |  |
| < 1.04mmol/L [men] & 1.3mmol/L [women] |  | 5,276 (18%) | 4,638 (17%) | 638 (41%) |  |
| Triglycerides [mean (SD)] | 31,440 | 1.64 (0.95) | 1.60 (0.92) | 2.24 (1.18) | <0.001 |
| Categories [n (%)] | 31,440 |  |  |  | <0.001 |
| ≤ 1.7 mmol/L |  | 20,301 (65%) | 19,664 (66%) | 627 (38%) |  |
| > 1.7 mmol/L |  | 11,139 (35%) | 10,115 (34%) | 1,034 (62%) |  |

|  | N | Whole cohort<br>(n=33,616) | cT1<800ms<br>(n=31,840) | cT1 ≥800ms<br>(n=1,776) | P-value<br>cT1<800ms<br>Vs<br>cT1≥800ms |
| --- | --- | --- | --- | --- | --- |
| **Total cholesterol (mmol/L)**<br>**[mean (SD)]** | 31,462 | 5.73 (1.08) | 5.74 (1.08) | 5.60 (1.17) | <0.001 |
| **Categories [n (%)]** | 31,462 |  |  |  | <0.001 |
| ≤ 5.2 mmol/L |  | 10,077 (32%) | 9,440 (32%) | 637 (38%) |  |
| > 5.2 mmol/L |  | 21,385 (68%) | 20,356 (68%) | 1,029 (62%) |  |
| **Any previous major CVD**<br>**event [n (%)]** | 33,610 | 4,111 (12%) | 3,782 (12%) | 329 (19%) | <0.001 |
| **Metabolic Syndrome [n (%)]** | 25,440 | 8,877 (35%) | 7,906 (33%) | 971 (70%) | <0.001 |
| **LIVER BIOMARKERS** |  |  |  |  |  |
| **ALT [mean (SD)]** | 31,458 | 23 (14) | 22 (13) | 33 (20) | <0.001 |
| **Categories [n (%)]** | 31,458 |  |  |  | <0.001 |
| ≤ 45 U/L |  | 30,299 (96%) | 28,877 (97%) | 1,422 (85%) |  |
| > 45 U/L |  | 1,159 (3.7%) | 917 (3.1%) | 242 (15%) |  |
| **AST [mean (SD)]** | 31,333 | 26 (10) | 25 (9) | 29 (12) | <0.001 |
| **Categories [n (%)]** | 31,333 |  |  |  | <0.001 |
| ≤ 40 U/L |  | 30,080 (96%) | 28,607 (96%) | 1,473 (89%) |  |
| > 40 U/L |  | 1,253 (4.0%) | 1,070 (3.6%) | 183 (11%) |  |
| **FIB4 [mean (SD)]** | 30,564 | 1.3 (0.58) | 1.3 (0.59) | 1.2 (0.53) | <0.001 |
| **Categories [n (%)]** | 30,564 |  |  |  | <0.001 |
| FIB4 < 1.3 |  | 17,962 (59%) | 16,875 (53%) | 1,087 (61%) |  |
| FIB4 ≥ 1.3 & < 2.67 |  | 12,107 (40%) | 11,597 (40%) | 510 (31%) |  |
| FIB4 ≥ 2.67 |  | 495 (2%) | 472 (1.6%) | 23 (1.4%) |  |
| **cT1 (ms) [mean (SD)]** | 33,616 | 700 (55) | 693 (44) | 840 (38) | <0.001 |
| **Categories [n (%)]** | 33,616 |  |  |  | - |
| < 800ms |  | 31,840 (95%) | - | - |  |
| ≥ 800ms |  | 1,776 (5.3%) | - | - |  |
| **PDFF (%) [mean (SD)]** | 33,616 | 4.9 (4.7) | 4.3 (3.6) | 15.9 (8.3) | <0.001 |
| **Categories [n (%)]** | 33,616 |  |  |  | <0.001 |
| < 5% |  | 24,503 (73%) | 24,269 (76%) | 234 (13%) |  |
| ≥ 5% |  | 9,113 (27%) | 7,571 (24%) | 1,542 (87%) |  |
| **CARDIAC BIOMARKERS** |  |  |  |  |  |
| **CRP (mg/mL) [mean (SD)]** | 31,387 | 2.01 (3.48) | 1.93 (3.41) | 3.51 (4.25) | <0.001 |
| **Categories [n (%)]** | 31,387 |  |  |  | <0.001 |
| ≤ 10 mg/L |  | 30,586 (97%) | 29,038 (98%) | 1,548 (93%) |  |
| > 10 mg/L |  | 801 (2.6%) | 690 (2.3%) | 111 (6.7%) |  |
| **NEW ONSET OUTCOMES [n (%)]** |  |  |  |  |  |
| **Any CVD event** | 29,499 | 794 (2.7%) | 734 (2.6%) | 60 (4.1%) | <0.001 |
| **CAD** | 31,582 | 409 (1.3%) | 380 (1.3%) | 29 (1.8%) | 0.066 |
| **MI** | 32,781 | 160 (0.5%) | 147 (0.5%) | 13 (0.8%) | 0.094 |
| **AF** | 32,536 | 352 (1.1%) | 321 (1.0%) | 31 (1.8%) | 0.002 |
| **VTE** | 32,787 | 77 (0.2%) | 73 (0.2%) | 4 (0.2%) | >0.9 |
| **HF** | 33,411 | 142 (0.4%) | 128 (0.4%) | 14 (0.8%) | 0.013 |
| **Stroke** | 32,936 | 157 (0.5%) | 146 (0.5%) | 11 (0.6%) | 0.3 |
| **CVD hospitalisation** | 33,610 | 1,267 (3.8%) | 1,145 (3.6%) | 122 (6.9%) | <0.001 |
| **All-cause mortality** | 33,610 | 236 (0.7%) | 210 (0.7%) | 26 (1.5%) | <0.001 |
| **Liver-related events** | 32,453 | 318 (0.9%) | 280 (0.9%) | 38 (2%) | <0.001 |

*Abbreviations: BMI, body mass index; HbA1C, glycated haemoglobin; ALT, alanine transaminase; AST, aspartate transaminase; LDL, low-density lipoprotein; HDL, high-density lipoprotein; CRP, c-reactive protein; cT1, iron-corrected T1 relaxation time; PDFF, proton density fat fraction; ANY CVD EVENT, Major adverse cardiovascular events; CAD, coronary artery disease; MI, myocardial infarction; AF, atrial fibrillation; VTE, Embolism/vascular event; HF, heart failure; CVD, cardiovascular disease.*

### Association between CVD outcomes and biomarkers treated as continuous variables

Elevation in liver cT1 was associated with a higher risk of all cardiovascular events investigated and all-cause mortality, in all cases independently of other liver biomarkers, including FIB4 and AST/ALT ratio **(Table 2)**. Smoking was not a significant as a univariable and therefore, was not included in the multivariable models (characteristics of the population with any major cardiovascular events can be found in **table S3**).

**Table 2.** Multivariate Analysis Cox Proportional Hazard Ratios with 95% Confidence Intervals of cardiovascular outcomes, based on Z-scores to enable comparisons across different unit scales. This table shows only HR from multivariable models, variables with “ns” were not included in these models.

| Metric | Outcome | HR | p value |
| --- | --- | --- | --- |
| cT1 (ms) | CVD Hospitalisation | 1.27 (1.18 - 1.37) | < 0.001 |
|  | Atrial Fibrillation | 1.3 (1.12 - 1.51) | < 0.001 |
|  | Heart Failure | 1.3 (1.08 - 1.58) | 0.004 |
|  | All-Cause Mortality | 1.19 (1.02 - 1.38) | 0.026 |
|  | Any CVD event | 1.14 (1.03 - 1.26) | 0.008 |
| PDFF (%) | CVD Hospitalisation | 0.87 (0.8 - 0.95) | 0.001 |
|  | Atrial Fibrillation | 0.91 (0.78 - 1.05) | 0.229 |
|  | Heart Failure | ns | ns |
|  | All-Cause Mortality | ns | ns |
|  | Any CVD event | 0.96 (0.87 - 1.06) | 0.402 |
| BMI (kg/m<sup>2</sup>) | CVD Hospitalisation | 1.05 (0.95 - 1.16) | 0.309 |
|  | Atrial Fibrillation | 1.14 (0.94 - 1.38) | 0.167 |
|  | Heart Failure | 0.87 (0.65 - 1.17) | 0.267 |
|  | All-Cause Mortality | 1.03 (0.81 - 1.29) | 0.825 |
|  | Any CVD event | 1.05 (0.92 - 1.19) | 0.47 |
| HbA1c (mmol/mol) | CVD Hospitalisation | 1.06 (1.01 - 1.12) | 0.011 |
|  | Atrial Fibrillation | 0.96 (0.86 - 1.08) | 0.554 |
|  | Heart Failure | 1.1 (0.97 - 1.25) | 0.086 |
|  | All-Cause Mortality | 1.06 (0.94 - 1.2) | 0.326 |
|  | Any CVD event | 1.04 (0.97 - 1.12) | 0.211 |
| Waist Circumference (cm) | CVD Hospitalisation | 1.19 (1.07 - 1.33) | 0.001 |
|  | Atrial Fibrillation | 1.17 (0.95 - 1.45) | 0.107 |
|  | Heart Failure | 1.43 (1.07 - 1.92) | 0.005 |
|  | All-Cause Mortality | 1.18 (0.92 - 1.53) | 0.182 |
|  | Any CVD event | 1.22 (1.06 - 1.4) | 0.003 |
| Total cholesterol | CVD Hospitalisation | 0.94 (0.89 - 1.01) | 0.079 |
|  | Atrial Fibrillation | 0.94 (0.83 - 1.06) | 0.28 |
|  | Heart Failure | 0.81 (0.67 - 0.98) | 0.017 |
|  | All-Cause Mortality | ns | ns |
|  | Any CVD event | ns | ns |
| AST | CVD Hospitalisation | 1.02 (0.91 - 1.15) | 0.616 |
|  | Atrial Fibrillation | 0.89 (0.68 - 1.16) | 0.347 |
|  | Heart Failure | 0.92 (0.73 - 1.16) | 0.483 |
|  | All-Cause Mortality | 0.98 (0.82 - 1.19) | 0.825 |
|  | Any CVD event | 0.94 (0.77 - 1.15) | 0.544 |
| ALT | CVD Hospitalisation | 0.99 (0.87 - 1.13) | 0.87 |
|  | Atrial Fibrillation | 1.03 (0.83 - 1.28) | 0.788 |
|  | Heart Failure | ns | ns |
|  | All-Cause Mortality | ns | ns |
|  | Any CVD event | 1.03 (0.85 - 1.24) | 0.779 |
| AST/ALT Ratio | CVD Hospitalisation | 1.01 (0.91 - 1.12) | 0.782 |
|  | Atrial Fibrillation | ns | ns |
|  | Heart Failure | ns | ns |
|  | All-Cause Mortality | ns | ns |
|  | Any CVD event | 0.99 (0.86 - 1.14) | 0.882 |
| C-Reactive Protein | CVD Hospitalisation | 1.02 (0.96 - 1.08) | 0.419 |
|  | Atrial Fibrillation | 0.99 (0.88 - 1.11) | 0.76 |
|  | Heart Failure | 1.1 (0.97 - 1.23) | 0.136 |
|  | All-Cause Mortality | ns | ns |
|  | Any CVD event | 1.03 (0.96 - 1.11) | 0.358 |
| FIB4 | CVD Hospitalisation | 0.97 (0.9 - 1.06) | 0.438 |
|  | Atrial Fibrillation | 1.01 (0.9 - 1.14) | 0.783 |
|  | Heart Failure | 1.06 (1.01 - 1.10) | 0.007 |
|  | All-Cause Mortality | 0.96 (0.8 - 1.15) | 0.56 |
|  | Any CVD event | 0.99 (0.9 - 1.09) | 0.788 |
| **Systolic Blood Pressure** | CVD Hospitalisation | 1.1 (1.03 - 1.17) | 0.004 |
|  | Atrial Fibrillation | 1.04 (0.92 - 1.17) | 0.573 |
|  | Heart Failure | 1.08 (0.89 - 1.3) | 0.432 |
|  | All-Cause Mortality | 0.94 (0.81 - 1.1) | 0.432 |
|  | Any CVD event | 1.1 (1.02 - 1.2) | 0.017 |
| **Age at MRI** | CVD Hospitalisation | 1.58 (1.46 - 1.71) | < 0.001 |
|  | Atrial Fibrillation | 1.95 (1.67 - 2.27) | < 0.001 |
|  | Heart Failure | 2.04 (1.61 - 2.59) | < 0.001 |
|  | All-Cause Mortality | 2.09 (1.72 - 2.54) | < 0.001 |
|  | Any CVD event | 1.69 (1.53 - 1.86) | < 0.001 |
| **Sex (Male)** | CVD Hospitalisation | 1.65 (1.39 - 1.95) | < 0.001 |
|  | Atrial Fibrillation | 1.79 (1.29 - 2.48) | < 0.001 |
|  | Heart Failure | 1.46 (0.89 - 2.4) | 0.125 |
|  | All-Cause Mortality | 1.33 (0.92 - 1.95) | 0.096 |
|  | Any CVD event | 1.62 (1.31 - 1.99) | < 0.001 |

*Abbreviations: BMI, body mass index; HbA1C, glycated haemoglobin; ALT, alanine transaminase; AST, aspartate transaminase; CRP, c-reactive protein; cT1, iron-corrected T1 relaxation time; PDFF, proton density fat fraction; Any CVD event, any cardiovascular event; CVD, cardiovascular; n.s., variables not significant in the univariate analysis were not included in the multivariate analysis.*

Cases where liver IDPs were not significantly associated with clinical outcomes (VTE, Stroke, MI and CAD) are described in **supplementary table S8**. Increasing liver cT1 was associated with higher risk of any CVD outcomes [HR(CI) 1.14(1.03-1.26), p=0.008] alongside waist circumference (p=0.003), SBP (p=0.017), age (p<0.001) and male sex (p<0.001) **(Table 2, Figure 2)**. Associations with specific CVD events are described below **(Table 2, supplementary table S8)**.

**Figure 2:**
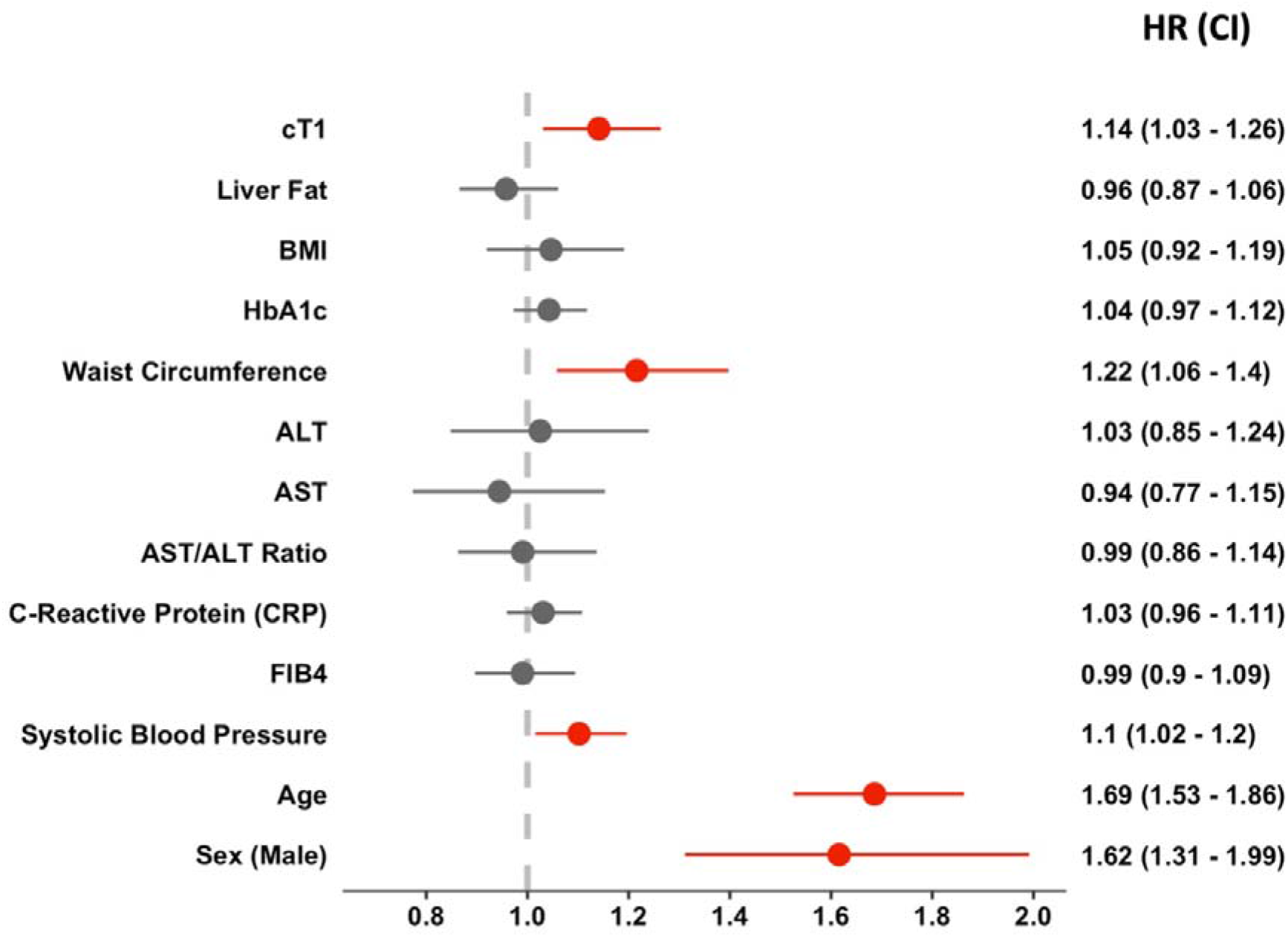
Multivariable model: Hazard ratios of associations for risk of any major CVD event in the whole cohort; variables treated as continuous based on Z-scores to enable comparisons across different unit scales. Significant associations shown in red.

#### Atrial Fibrillation

AF was significantly associated with increasing liver cT1 [HR(CI) 1.30 (1.12-1.51), p<0.001], age (p<0.001) and male sex (p<0.001), (characteristics of the population with atrial fibrillation events can be found in **table S4**)

#### Heart Failure

HF was significantly associated with increasing liver cT1 [HR(CI)1.30 (1.09-1.56), p=0.004] alongside with waist circumference (p=0.005), FIB4 (p=0.007) and age (p<0.001), while total cholesterol was negatively associated (p=0.017), (characteristics of the population with heart failure events can be found in **table S5**)

#### Hospitalisation due to cardiovascular events

Higher risk of hospitalisation from cardiovascular causes was associated with increasing liver cT1 [HR(CI) 1.27(1.18-1.37), p<0.001] and higher HbA1c [HR(CI)1.06 (1.01-1.12), p=0.011] as well as waist circumference (p=0.001), SBP (p=0.004), age (p<0.001) and male sex (p<0.001), (characteristics of the population being hospitalised due to CVD events can be found in **table S6**)

#### All-cause mortality

Higher liver cT1 [HR(CI) 1.19(1.02-1.38), p=0.026], and age (p<0.001) were significantly associated with all-cause mortality, (characteristics of the population deceased after MRI scan can be found in **table S7**)

### Association between CVD outcomes and biomarkers using clinically used thresholds

Associations to CVD events were assessed using validated clinical biomarker thresholds and demographic categories. cT1 threshold was _≥_800ms as this is considered the upper limit of normal and the recommended threshold to identify those in transition from simple steatosis to NASH (28). The threshold for elevated liver fat was PDFF_≥_ 5% and for elevated fibrosis was FIB4 index >1.3 and _≥_2.67 (based on established guidelines (11,28,32)(39), respectively).

N=1776 individuals had cT1 _≥_800ms (Table 1); of whom 70% had metabolic syndrome and 87% clinically significant liver steatosis (PDFF_≥_5%). Participants with cT1_≥_800ms experienced a two-fold higher number of cardiovascular events (N=122, 7%) compared to those with values of cT1 <800ms (N=1145, 4%) **(Figure 3)**.

**Figure 3:**
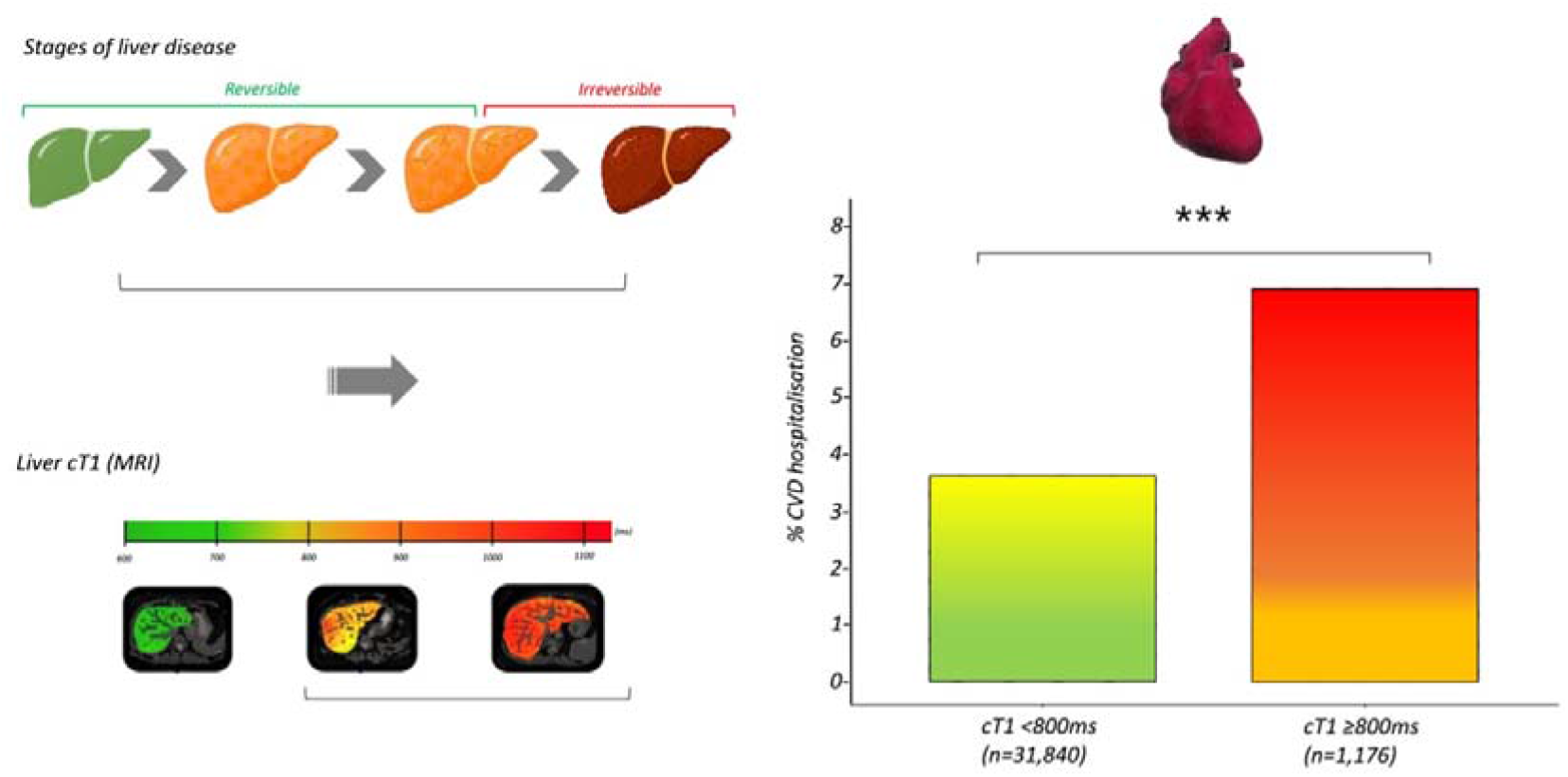
Pattern of liver disease progression (top left) and exemplar liver cT1 images for advancing liver disease (bottom left). Pattern of CVD increased hospitalisation events in those with evidence of liver disease as evidence from a cT1 above the upper limit of normal (right); *** represents P<.001

In the multivariable models, cT1 _≥_800ms was significantly associated with the risk of hospitalisation due to cardiovascular disease [HR (CI): 1.38 (1.11-1.75) p=0.005], other liver biomarkers were not: PDFF _≥_5% (p=0.5), ALT >45U/L in presence of diabetes and >50 U/L in absence of diabetes (p=0.64); AST >40U/L (p=0.43); AST/ALT ratio (p=0.87) and FIB4 (_≥_1.3 <2.67, p=0.69; _≥_2.67 points, p=0.87)]. Other exposures associated with CVD hospitalisation were BMI _≥_25kg/m2 (p=0.016) or BMI _≥_30kg/m2 (p<0.001), diabetes (HbA1c _≥_48mmol/mol) (p=0.028), older age (p<0.001) and male sex (p<0.001). Other known CVD risk factors such as total cholesterol _≥_5.2 mmol/L (p=0.275) and hypertension (p=0.065) were not significantly associated with CVD hospitalisation in this subgroup **(Figure 4)**.

**Figure 4:**
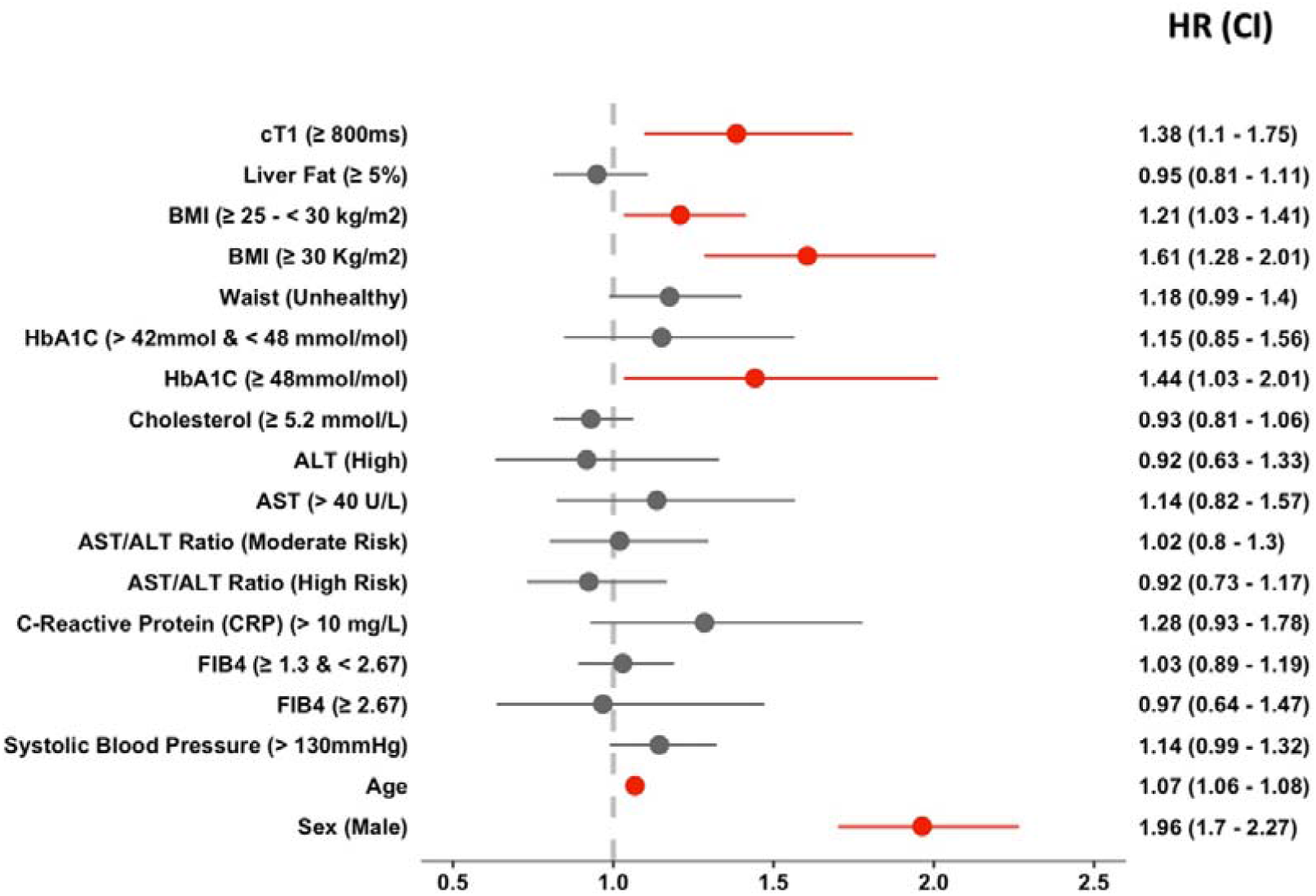
Hazard ratios of associations for risk of CVD hospitalisation in the whole cohort; variables treated as binary based on pre-specified clinical thresholds. Note: 95% CI for age effect is too small to be appreciated in the plot. Significant associations shown in red.

### CVD hospitalisation risk in individuals without metabolic disease

In this group of 16,563 individuals, characterised by lower age [mean 64 (7.6) years, p<0.001], lower BMI [mean 25 (3.8) kg/m^2^, p<0.001] and lower prevalence of clinically significant liver fat (17%, p<0.001), liver cT1 was associated with CVD hospitalisation [HR(CI) 1.26(1.13-1.4), p<0.001]. No other liver biomarkers showed any association in this subset. Age and male sex were associated with all CVD events, SBP with any major CVD event and HbA1c with CVD hospitalisations (**Supplementary table S9)**. All-cause mortality was associated with age only.

## DISCUSSION

In this large-scale, longitudinal study of cardiovascular disease event incidence in a mainly healthy older population, we have identified specific associations between individual liver biomarkers and CVD events. We found that, firstly, elevation of liver cT1, a liver specific marker of disease activity, was associated with increased risk of new onset for any CVD, and specifically AF, HF, CVD hospitalisation and all-cause mortality. In contrast, the commonly used liver risk score FIB4 had a much lesser predictive value, being associated with HF only, while the AST/ALT ratio was not predictive of any adverse events. Secondly, we identified that for clinical events of a higher prevalence (such as CVD hospitalisation), only cT1 at or above the clinically defined threshold that is used to diagnose CLD activity, remained associated; non-invasive blood screening tests for liver fibrosis were not. Thirdly, in a sensitivity analysis to remove those with pre-existing metabolic syndrome, the independent association between elevated cT1 and increased incident CVD hospitalisation remained, ç. These results highlight the prognostic relevance of a comprehensive evaluation of liver health in populations at risk of cardiovascular and/or CLD, even in the absence of clinical manifestations or metabolic syndrome, when there is an opportunity to modify/address risk factors and prevent disease progression. Given the recognition of T1 mapping in clinical guidelines for cardiac health as well as liver health, these results are highlighting the opportunity for quantitative imaging-based measurements to play a key role in shared cardio-metabolic pathways.

Prior studies have observed liver-related clinical events in NAFLD linked to the stage/severity of fibrosis, as measured from blood-based ELF test (40) or using other imaging techniques e.g. liver stiffness by magnetic resonance elastography (MRE) (41) or transient elastography from ultrasound. Using the current MR-based technique, cT1 has also been associated with all-cause mortality and event-free survival in patients with CLD(4). Whilst prognostic (fibrosis-based) assessment of risk in those with CLD may be of use in the liver clinic, it may be argued that this is rather late in the disease course. Measurements of liver health that allow for risk stratification across the spectrum of early to late stages have the potential to be transformative in terms of personalized care. The results observed in this study showed a robust link between evidence of measurable CLD activity change from cT1 and a variety of cardiovascular events. Of clinical relevance is the link between cT1_≥_800ms and hospitalisation for CVD. Previous literature has reported cT1_≥_800ms to have excellent diagnostic accuracy for identifying patients with NASH from healthy participants (28). The combination of these findings has important clinical implications as it enables the distinction of patients with clinically significant steatohepatitis at an early and modifiable stage of CLD from those with more benign simple steatosis.

Significantly, another defining feature of NAFLD, accumulation of liver fat (steatosis), although univariately associated with various CVD outcomes, was not independently associated with any of the clinical outcomes in multivariable analyses. On one occasion lower PDFF appeared to be significantly associated with CVD hospitalisation. This result is interesting, and in this instance, given the linear relationship observed when included as a univariate analysis, it is most likely an indicator of the multicollinearity observed between PDFF and other variables. It should be noted, however, that lower PDFF is a common feature of advanced CLD and cirrhosis and therefore is not a biomarker that can reliably be used for risk stratification. There was also no association between the blood-based algorithm for cirrhosis risk, the AST/ALT ratio, and CVD outcomes, and the FIB4 index was only observed to be associated with higher incidence of HF in our analysis. While previous work in the Rotterdam general population cohort study has shown that liver fibrosis, evaluated by transient elastography, has been associated with 7% AF prevalence, no association between incident AF and fibrosis was described (42) and no independent association seen with FIB4 in a CAD cohort (43). This, together with our findings suggesting that the link between CVD risk in the general population associated with liver health is likely related to underlying disease activity and not fibrosis, supports previous hypotheses of the underlying mechanisms related to tissue inflammation and metabolic processes.

Whilst there are no approved therapeutic agents yet in NAFLD, agents such as the glucagon-like peptide-1 receptor agonist (GLP-1), semaglutide, have been incorporated into clinical guidelines in those with type 2 diabetes as a treatment for non-alcoholic steatohepatitis (NASH), the aggressive form of NAFLD (10,44). In addition, tirzepatide (a dual glucose-dependent insulinotropic polypeptide (GIP)/GLP-1 agonist), which has been approved for weight loss in those with type 2 diabetes, has also recently been shown to achieve meaningful and sustained weight reduction in non-diabetic obese patients (45) and reductions in liver fat (46). These positive effects on metabolic health may also improve liver-related health, thus potentially having a profound modifiable effect on CVD risk. Markers for early-stage liver disease, such as cT1, may be considered as non-invasive alternatives to biopsy to monitor response and personalize treatments. cT1 has already been shown as an effective pharmaco-dynamic biomarker in NASH trials (7,8), and being inherently non-invasive, is an attractive tool for assessing early response in drug development. The current results showing a robust link to clinical outcomes, coupled with response to therapy, are suggestive of a place for cT1 in future NASH trials as a surrogate endpoint.

Many CVD risk scores exist, including the QRISK score, Framingham score and ASCVD score, which are already employed clinically. However, considering these results, and the momentum towards appreciating multi-system disease and multi-speciality care, our results highlight an opportunity to improve on these risk scores by incorporating the degree of liver-related disease activity. In relation to the FIB4 index, whilst we did not observe a robust association with CVD risk, it should be acknowledged that the currently adopted thresholds to rule out or rule in significant liver fibrosis are designed for patients being specifically evaluated for CLD (10,47) and may be inappropriate in ‘healthier’ populations (48) where CLD is underdiagnosed or at an earlier, potentially more modifiable, stage (49). Of course, the fact that we did not observe association with the FIB4 index is being attributed to the likely absence of fibrosis, but a notable limitation of the UK Biobank imaging study is that there is a delay of a number of years between the blood tests and the imaging which may prevent meaningful interpretation of the blood test results. Other notable limitations are the lack of confirmatory biopsy, although in following the guiding medical principal of *primum non nocere* this is not surprising in a study of the general population. The study cohort is also homogenous with a predominant white ethnicity. Despite these limitations, the results of our study reinforce the utility of cT1 in evaluating cardiometabolic risk in patients, highlighting cT1 as a prognostic non-invasive imaging biomarker that can stratify patients for therapy.

## CONCLUSION

In a population-based study we observed CVD events in 4% of people which were independently associated with evidence of CLD. These results suggest the MRI-derived biomarker cT1 has a promising role to play in risk stratification of those at greatest risk of CVD morbidity and mortality.

## Data Availability

Data Availability Statement
The data analyzed in this study is subject to the following licenses/restrictions: Data belongs to UK Biobank. Requests to access these datasets should be directed to.

https://www.ukbiobank.ac.uk/

## ACKNOWLEGDEMENTS

SN acknowledges support from the Oxford NIHR Biomedical Research Centre.

BR is funded by the British Heart Foundation Oxford Centre of Research Excellence (RE/18/3/34214)

## ETHICS STATEMENT

The studies involving human participants were reviewed and approved by 11/NW/0382. The patients/participants provided their written informed consent to participate in this study.

