## Supplementary file for "Liver disease is a significant risk factor for cardiovascular outcomes - a UK Biobank study"

**S1 Table:** **ICD-10 codes used for the definition of the CVD event category and individual sub-categories extracted from HESIN data in the UK Biobank**.

| **ICD category** | **ICD subcategory** |
| --- | --- |
| **ANY CVD EVENT (contains all ICD-10 below)** | |
| **Coronary artery disease (CAD) – includes MI** | |
| I24 Other acute ischaemic heart diseases | I24.0 Coronary thrombosis not resulting in myocardial infarction |
| I24 Other acute ischaemic heart diseases | I24.1 Dressler's syndrome |
| I25 Chronic ischaemic heart disease | I25.5 Ischaemic cardiomyopathy |
| I25 Chronic ischaemic heart disease | I25.6 Silent myocardial ischaemia |
| I25 Chronic ischaemic heart disease | I25.8 Other forms of chronic ischaemic heart disease |
| I25 Chronic ischaemic heart disease | I25.9 Chronic ischaemic heart disease, unspecified |
| I20 Angina pectoris | I20.0 Unstable angina |
| I20 Angina pectoris | I20.1 Angina pectoris with documented spasm |
| I20 Angina pectoris | I20.8 Other forms of angina pectoris |
| I20 Angina pectoris | I20.9 Angina pectoris, unspecified |
| **Myocardial infarction (MI)** | |
| I21 Acute myocardial infarction | I21.0 Acute transmural myocardial infarction of anterior wall |
| I21 Acute myocardial infarction | I21.1 Acute transmural myocardial infarction of inferior wall |
| I21 Acute myocardial infarction | I21.2 Acute transmural myocardial infarction of other sites |
| I21 Acute myocardial infarction | I21.3 Acute transmural myocardial infarction of unspecified site |
| I21 Acute myocardial infarction | I21.4 Acute subendocardial myocardial infarction |
| I21 Acute myocardial infarction | I21.9 Acute myocardial infarction, unspecified |
| I22 Subsequent myocardial infarction | I22.0 Subsequent myocardial infarction of anterior wall |
| I22 Subsequent myocardial infarction | I22.1 Subsequent myocardial infarction of inferior wall |
| I22 Subsequent myocardial infarction | I22.8 Subsequent myocardial infarction of other sites |
| I22 Subsequent myocardial infarction | I22.9 Subsequent myocardial infarction of unspecified site |
| I23 Certain current complications following acute myocardial infarction | I23.0 Haemopericardium as current complication following acute myocardial infarction |
| I23 Certain current complications following acute myocardial infarction | I23.1 Atrial septal defect as current complication following acute myocardial infarction |
| I23 Certain current complications following acute myocardial infarction | I23.2 Ventricular septal defect as current complication following acute myocardial infarction |
| I23 Certain current complications following acute myocardial infarction | I23.3 Rupture of cardiac wall without haemopericardium as current complication following acute myocardial infarction |
| I23 Certain current complications following acute myocardial infarction | I23.4 Rupture of chordae tendineae as current complication following acute myocardial infarction |
| I23 Certain current complications following acute myocardial infarction | I23.5 Rupture of papillary muscle as current complication following acute myocardial infarction |
| I23 Certain current complications following acute myocardial infarction | I23.6 Thrombosis of atrium, auricular appendage and ventricle as current complications following acute myocardial infarction |
| I23 Certain current complications following acute myocardial infarction | I23.8 Other current complications following acute myocardial infarction |
| **Atrial fibrillation (AF)** | |
| I48 Atrial fibrillation and flutter | I48.0 Paroxysmal atrial fibrillation |
| I48 Atrial fibrillation and flutter | I48.1 Persistent atrial fibrillation |
| I48 Atrial fibrillation and flutter | I48.2 Chronic atrial fibrillation |
| I48 Atrial fibrillation and flutter | I48.9 Atrial fibrillation and atrial flutter, unspecified |
| **Embolism/vascular event (VTE)** | |
| I74 Arterial embolism and thrombosis | I74.0 Embolism and thrombosis of abdominal aorta |
| I74 Arterial embolism and thrombosis | I74.1 Embolism and thrombosis of other and unspecified parts of aorta |
| I74 Arterial embolism and thrombosis | I74.2 Embolism and thrombosis of arteries of the upper extremities |
| I74 Arterial embolism and thrombosis | I74.3 Embolism and thrombosis of arteries of the lower extremities |
| I74 Arterial embolism and thrombosis | I74.4 Embolism and thrombosis of arteries of extremities, unspecified |
| I74 Arterial embolism and thrombosis | I74.5 Embolism and thrombosis of iliac artery |
| I74 Arterial embolism and thrombosis | I74.8 Embolism and thrombosis of other arteries |
| I74 Arterial embolism and thrombosis | I74.9 Embolism and thrombosis of unspecified artery |
| I80 Phlebitis and thrombophlebitis | I80.0 Phlebitis and thrombophlebitis of superficial vessels of lower extremities |
| I80 Phlebitis and thrombophlebitis | I80.1 Phlebitis and thrombophlebitis of femoral vein |
| I80 Phlebitis and thrombophlebitis | I80.2 Phlebitis and thrombophlebitis of other deep vessels of lower extremities |
| I80 Phlebitis and thrombophlebitis | I80.3 Phlebitis and thrombophlebitis of lower extremities, unspecified |
| I80 Phlebitis and thrombophlebitis | I80.8 Phlebitis and thrombophlebitis of other sites |
| I80 Phlebitis and thrombophlebitis | I80.9 Phlebitis and thrombophlebitis of unspecified site |
| I82 Other venous embolism and thrombosis | I82.2 Embolism and thrombosis of vena cava |
| I82 Other venous embolism and thrombosis | I82.3 Embolism and thrombosis of renal vein |
| I82 Other venous embolism and thrombosis | I82.8 Embolism and thrombosis of other specified veins |
| I82 Other venous embolism and thrombosis | I82.9 Embolism and thrombosis of unspecified vein |
| **Heart failure (HF)** | |
| I50 Heart failure | I50.0 Congestive heart failure |
| I50 Heart failure | I50.1 Left ventricular failure |
| I50 Heart failure | I50.9 Heart failure, unspecified |
| **Stroke** | |
| I60 Subarachnoid haemorrhage | I60.0 Subarachnoid haemorrhage from carotid siphon and bifurcation |
| I60 Subarachnoid haemorrhage | I60.1 Subarachnoid haemorrhage from middle cerebral artery |
| I60 Subarachnoid haemorrhage | I60.2 Subarachnoid haemorrhage from anterior communicating artery |
| I60 Subarachnoid haemorrhage | I60.3 Subarachnoid haemorrhage from posterior communicating artery |
| I60 Subarachnoid haemorrhage | I60.4 Subarachnoid haemorrhage from basilar artery |
| I60 Subarachnoid haemorrhage | I60.5 Subarachnoid haemorrhage from vertebral artery |
| I60 Subarachnoid haemorrhage | I60.6 Subarachnoid haemorrhage from other intracranial arteries |
| I60 Subarachnoid haemorrhage | I60.7 Subarachnoid haemorrhage from intracranial artery, unspecified |
| I60 Subarachnoid haemorrhage | I60.8 Other subarachnoid haemorrhage |
| I60 Subarachnoid haemorrhage | I60.9 Subarachnoid haemorrhage, unspecified |
| I61 Intracerebral haemorrhage | I61.0 Intracerebral haemorrhage in hemisphere, subcortical |
| I61 Intracerebral haemorrhage | I61.1 Intracerebral haemorrhage in hemisphere, cortical |
| I61 Intracerebral haemorrhage | I61.2 Intracerebral haemorrhage in hemisphere, unspecified |
| I61 Intracerebral haemorrhage | I61.3 Intracerebral haemorrhage in brain stem |
| I61 Intracerebral haemorrhage | I61.4 Intracerebral haemorrhage in cerebellum |
| I61 Intracerebral haemorrhage | I61.5 Intracerebral haemorrhage, intraventricular |
| I61 Intracerebral haemorrhage | I61.6 Intracerebral haemorrhage, multiple localised |
| I61 Intracerebral haemorrhage | I61.8 Other intracerebral haemorrhage |
| I61 Intracerebral haemorrhage | I61.9 Intracerebral haemorrhage, unspecified |
| I62 Other nontraumatic intracranial haemorrhage | I62.0 Subdural haemorrhage (acute) (nontraumatic) |
| I62 Other nontraumatic intracranial haemorrhage | I62.1 Nontraumatic extradural haemorrhage |
| I62 Other nontraumatic intracranial haemorrhage | I62.9 Intracranial haemorrhage (nontraumatic), unspecified |
| I63 Cerebral infarction | I63.0 Cerebral infarction due to thrombosis of precerebral arteries |
| I63 Cerebral infarction | I63.1 Cerebral infarction due to embolism of precerebral arteries |
| I63 Cerebral infarction | I63.2 Cerebral infarction due to unspecified occlusion or stenosis of precerebral arteries |
| I63 Cerebral infarction | I63.3 Cerebral infarction due to thrombosis of cerebral arteries |
| I63 Cerebral infarction | I63.4 Cerebral infarction due to embolism of cerebral arteries |
| I63 Cerebral infarction | I63.5 Cerebral infarction due to unspecified occlusion or stenosis of cerebral arteries |
| I63 Cerebral infarction | I63.6 Cerebral infarction due to cerebral venous thrombosis, nonpyogenic |
| I63 Cerebral infarction | I63.8 Other cerebral infarction |
| I63 Cerebral infarction | I63.9 Cerebral infarction, unspecified |
| I64 Stroke, not specified as haemorrhage or infarction | I64 Stroke, not specified as haemorrhage or infarction |
| I69 Sequelae of cerebrovascular disease | I69.0 Sequelae of subarachnoid haemorrhage |
| I69 Sequelae of cerebrovascular disease | I69.1 Sequelae of intracerebral haemorrhage |
| I69 Sequelae of cerebrovascular disease | I69.2 Sequelae of other nontraumatic intracranial haemorrhage |
| I69 Sequelae of cerebrovascular disease | I69.3 Sequelae of cerebral infarction |
| I69 Sequelae of cerebrovascular disease | I69.4 Sequelae of stroke, not specified as haemorrhage or infarction |
| I69 Sequelae of cerebrovascular disease | I69.8 Sequelae of other and unspecified cerebrovascular diseases |

The HESIN category contains data on hospital inpatient admissions, obtained through linkage to external data providers. Inpatients are defined as persons who are admitted to hospital and occupy a hospital bed. This includes both admissions where an overnight stay is planned and day cases.

**S2: Liver imaging protocol*:**

| **Sequence** | **Acquisition parameters  (as reported by (1))** | **Processing, quality control and analysis** | **Imaging-derived phenotypes produced** | **Key references for full protocol** |
| --- | --- | --- | --- | --- |
| shMOLLI  (shortened modified Look-Locker imaging) | Resolution: 1.146 x 1.146 x 8,  Matrix: 384 x 288 x 7  TE/TR = 1.93/480.6 ms, α = 35°, R = 2 | Main extraction and analysis via Perspectum’s LiverMultiScan™ Discover software (2)  PDFF maps were constructed, using a three-point DIXON technique (3)  Further large-scale phenotyping via a deep learning method (4) | Proton density fat fraction (PDFF)  Liver iron  Corrected t1 (ct1) | McKay et al. (2018)(5)  Mojtahed et al. (2019)(6)  Wilman et al. (2017)(7) Perpectum (2016) (8) |
| LMS LiverMultiScan | Resolution: 1.719 x 1.719 x 10  Matrix: 256 x 232 x 6  TE/TR = 1.2 (min) TE/TR = 7.2 (max)/14 ms, α = 5° |  |  |  |
| ME GRE  (multi-echo spoiled-gradient-echo acquisition) | Resolution: 2.5 x 2.5 x 6  Matrix: 160 x 160 x 10  TE/TR = 2.38(min) TE/TR = 23.8(max)/27 ms, α = 20° |  |  |  |

*Scans have been acquired on Siemens 1.5T MAGNETOM Aera.

**S3: Table:** Characteristics of participants with any major cardiovascular events:

|  | | **Any major CVD event** | | |
| --- | --- | --- | --- | --- |
|  | | No major CVD event (N=28,705) | Major CVD event (n=794) | P Value |
| **Demographics** | | | | |
| Age [mean (SD)] | | 64 (8) | 67 (7) | **<0.001** |
| Sex (Male) [n (%)] | | 12,783 (45%) | 505 (64%) | **<0.001** |
| Race (White British) [n (%)] | | 26,066 (91%) | 732 (92%) | 0.19 |
| Smoking [n (%)] | | 911 (3.2%) | 26 (3.3%) | 0.87 |
| **Metabolic comorbidities** | | | | |
| BMI at MRI [mean (SD)] | | 26.2 (4.2) | 27.1 (4.3) | **<0.001** |
| Categories [n (%)] | |  |  | **<0.001** |
| Lean (BMI < 25Kg/m^2^) | | 12,423 (43%) | 270 (34%) |  |
| Overweight (BMI ≥ 25 - < 30 kg/m^2^) | | 11,627 (41%) | 343 (43%) |  |
| Obese (BMI ≥ 30 Kg/m^2^) | | 4,654 (16%) | 181 (23%) |  |
| Systolic blood pressure (SBP, mmHg) [mean (SD)] | | 136 (19) | 143 (19) | **<0.001** |
| Categories [n (%)] | |  |  | **<0.001** |
| SBP ≤ 130mmHg | | 11,089 (41%) | 207 (28%) |  |
| SBP > 130mmHg | | 15,665 (59%) | 542 (72%) |  |
| HbA1C (mmol/mol) [mean (SD)] | | 34.8 (5.0) | 36.1 (6.1) | **<0.001** |
| Categories [n (%)] | |  |  | **<0.001** |
| ≤ 42mmol/mol | | 25,623 (96%) | 683 (93%) |  |
| 42mmol & < 48 mmol/mol | | 594 (2.2%) | 30 (4.1%) |  |
| ≥ 48mmol/mol | | 431 (1.6%) | 21 (2.9%) |  |
| LDL [mean (SD)] | | 3.60 (0.82) | 3.65 (0.83) | 0.10 |
| Categories [n (%)] | |  |  | **0.02** |
| ≤ 4.1 mmol/L | | 19,836 (74%) | 516 (70%) |  |
| > 4.1 mmol/L | | 6,978 (26%) | 218 (30%) |  |
| HDL [mean (SD)] | | 1.50 (0.38) | 1.39 (0.36) | **<0.001** |
| Categories [n (%)] | |  |  | **0.003** |
| ≥ 1.04mmol/L [men] & 1.3mmol/L [women] | | 20,294 (83%) | 513 (78%) |  |
| < 1.04mmol/L [men] & 1.3mmol/L [women] | | 4,271 (17%) | 143 (22%) |  |
| Triglycerides [mean (SD)] | | 1.61 (0.93) | 1.82 (0.95) | **<0.001** |
| Categories [n (%)] | | |  | **<0.001** |
| ≤ 1.7 mmol/L | | 17,731 (66%) | 413 (56%) |  |
| > 1.7 mmol/L | | 9,124 (34%) | 322 (44%) |  |
| Total cholesterol (mmol/L) [mean (SD)] | | 5.77 (1.06) | 5.77 (1.09) | 0.94 |
| Categories [n (%)] | | |  | 0.21 |
| ≤ 5.2 mmol/L | | 8,246 (31%) | 210 (29%) |  |
| > 5.2 mmol/L | | 18,626 (69%) | 526 (71%) |  |
| Metabolic Syndrome [n (%)] | | 7,135 (33%) | 285 (48%) | **<0.001** |
| **Liver biomarkers** | | | | |
| ALT [mean (SD)] | | |  | **<0.001** |
| Categories [n (%)] | 23 (14) | | 25 (14) | 0.10 |
| ≤ 45 U/L | 25,927 (96%) | | 702 (95%) |  |
| > 45 U/L | 943 (3.5%) | | 34 (4.6%) |  |
| AST [mean (SD)] | 25 (9) | | 26 (8) | **0.0009** |
| Categories [n (%)] |  | |  | 0.10 |
| ≤ 40 U/L | 25,756 (96%) | | 699 (95%) |  |
| > 40 U/L | 1,003 (3.7%) | | 36 (4.9%) |  |
| FIB4 [mean (SD)] | 1.27 (0.58) | | 1.38 (0.51) | **<0.001** |
| Categories [n (%)] |  | |  | **<0.001** |
| FIB4 < 1.3 | 15,882 (61%) | | 367 (51%) |  |
| FIB4 ≥ 1.3 & < 2.67 | 9,839 (38%) | | 335 (47%) |  |
| FIB4 ≥ 2.67 | 369 (1.4%) | | 18 (2.5%) |  |
| cT1 (ms) [mean (SD)] | 698 (54) | | 712 (57) | **<0.001** |
| Categories [n (%)] |  | |  | **<0.001** |
| < 800ms | 27,318 (95%) | | 734 (92%) |  |
| ≥ 800ms | 1,387 (4.8%) | | 60 (7.6%) |  |
| PDFF (%) [mean (SD)] | 4.8 (4.7) | | 5.6 (5.2) | **<0.001** |
| Categories [n (%)] |  | |  | **<0.001** |
| < 5% | 21,126 (74%) | | 522 (66%) |  |
| ≥ 5% | 7,579 (26%) | | 272 (34%) |  |
| **Cardiac biomarkers** | | | | |
| CRP (mg/mL) [mean (SD)] | | 1.96 (3.41) | 2.35 (3.59) | **0.004** |
| Categories [n (%)] | |  |  | 0.09 |
| ≤ 10 mg/L | | 26,162 (98%) | 705 (97%) |  |
| > 10 mg/L | | 656 (2.4%) | 25 (3.4%) |  |

**S4 Table:** Characteristics of participants with new onset atrial fibrillation:

|  | **Atrial Fibrillation** | | |
| --- | --- | --- | --- |
|  | No AF (n=32,184) | AF (n=352) | P Value |
| **Demographics** | |  |  |
| Age [mean (SD)] | 64 (8) | 68 (6) | **<0.001** |
| Sex (Male) [n (%)] | 14,947 (46%) | 237 (67%) | **<0.001** |
| Race (White British) [n (%)] | 29,245 (91%) | 322 (91%) | 0.70 |
| Smoking [n (%)] | 1,031 (3.2%) | 11 (3.1%) | 0.93 |
| **Metabolic comorbidities** | |  |  |
| BMI at MRI [mean (SD)] | 26.3 (4.2) | 27.5 (4.4) | **<0.001** |
| Categories [n (%)] |  |  | **<0.001** |
| Lean (BMI < 25Kg/m^2^) | 13,503 (42%) | 101 (29%) |  |
| Overweight (BMI ≥ 25 - < 30 kg/m^2^) | 13,216 (41%) | 158 (45%) |  |
| Obese (BMI ≥ 30 Kg/m^2^) | 5,464 (17%) | 93 (26%) |  |
| Systolic blood pressure (SBP, mmHg) [mean (SD)] | 136 (19) | 143 (19) | **<0.001** |
| Categories [n (%)] |  |  | **<0.001** |
| SBP ≤ 130mmHg | 12,128 (40%) | 95 (29%) |  |
| SBP > 130mmHg | 17,874 (60%) | 235 (71%) |  |
| HbA1C (mmol/mol) [mean (SD)] | 35.0 (5.2) | 36.1 (6.8) | **0.004** |
| Categories [n (%)] |  |  | 0.052 |
| ≤ 42mmol/mol | 28,611 (96%) | 301 (93%) |  |
| 42mmol & < 48 mmol/mol | 750 (2.5%) | 11 (3.4%) |  |
| ≥ 48mmol/mol | 534 (1.8%) | 11 (3.4%) |  |
| LDL [mean (SD)] | 3.59 (0.83) | 3.50 (0.84) | **0.04** |
| Categories [n (%)] |  |  | 0.11 |
| ≤ 4.1 mmol/L | 22,250 (74%) | 253 (78%) |  |
| > 4.1 mmol/L | 7,811 (26%) | 72 (22%) |  |
| HDL [mean (SD)] | 1.48 (0.38) | 1.39 (0.37) | **<0.001** |
| Categories [n (%)] |  |  | **0.012** |
| ≥ 1.04mmol/L [men] & 1.3mmol/L [women] | 22,495 (82%) | 223 (76%) |  |
| < 1.04mmol/L [men] & 1.3mmol/L [women] | 5,006 (18%) | 70 (24%) |  |
| Triglycerides [mean (SD)] | 1.63 (0.95) | 1.81 (0.95) | **0.001** |
| Categories [n (%)] | |  | **0.018** |
| ≤ 1.7 mmol/L | 19,495 (65%) | 190 (58%) |  |
| > 1.7 mmol/L | 10,609 (35%) | 135 (42%) |  |
| Total cholesterol (mmol/L) [mean (SD)] | 5.74 (1.08) | 5.58 (1.11) | **0.008** |
| Categories [n (%)] | |  | **0.18** |
| ≤ 5.2 mmol/L | 9,523 (32%) | 114 (35%) |  |
| > 5.2 mmol/L | 20,601 (68%) | 211 (65%) |  |
| Metabolic Syndrome [n (%)] | 8,394 (34%) | 130 (49%) | **<0.001** |
| **Liver biomarkers** | |  |  |
| ALT [mean (SD)] | 23 (14) | 25 (13) | **0.006** |
| Categories [n (%)] |  |  | 0.53 |
| ≤ 45 U/L | 29,019 (96%) | 311 (96%) |  |
| > 45 U/L | 1,101 (3.7%) | 14 (4.3%) |  |
| AST [mean (SD)] | 26 (10) | 27 (7) | **0.017** |
| Categories [n (%)] |  |  | 0.07 |
| ≤ 40 U/L | 28,826 (96%) | 306 (94%) |  |
| > 40 U/L | 1,173 (3.9%) | 19 (5.8%) |  |
| FIB4 [mean (SD)] | 1.29 (0.58) | 1.45 (0.56) | **<0.001** |
| Categories [n (%)] |  |  | **<0.001** |
| FIB4 < 1.3 | 17,423 (60%) | 152 (47%) |  |
| FIB4 ≥ 1.3 & < 2.67 | 11,398 (39%) | 156 (49%) |  |
| FIB4 ≥ 2.67 | 436 (1.5%) | 13 (4.0%) |  |
| cT1 (ms) [mean (SD)] | 700 (55) | 719 (58) | **<0.001** |
| Categories [n (%)] | |  | **0.001** |
| < 800ms | 30,534 (95%) | 321 (91%) |  |
| ≥ 800ms | 1,650 (5.1%) | 31 (8.8%) |  |
| PDFF (%) [mean (SD)] | 4.9 (4.7) | 5.8 (5.5) | **0.002** |
| Categories [n (%)] |  |  | **0.001** |
| < 5% | 23,476 (73%) | 230 (65%) |  |
| ≥ 5% | 8,708 (27%) | 122 (35%) |  |
| **Cardiac biomarkers** | |  |  |
| CRP (mg/mL) [mean (SD)] | 2.00 (3.48) | 2.30 (2.84) | 0.06 |
| Categories [n (%)] |  |  |  |
| ≤ 10 mg/L | 29,295 (97%) | 313 (98%) | 0.95 |
| > 10 mg/L | 763 (2.5%) | 8 (2.5%) |  |

**S5 Table:** Characteristics of participants with new onset heart failure:

|  | **Heart Failure** | | |
| --- | --- | --- | --- |
|  | No HF (n=33,269) | HF (n=142) | P Value |
| **Demographics** | |  |  |
| Age [mean (SD)] | 64 (8) | 69 (6) | **<0.001** |
| Sex (Male) [n (%)] | 15,683 (47%) | 100 (70%) | **<0.001** |
| Race (White British) [n (%)] | 30,255 (91%) | 137 (96%) | **0.02** |
| Smoking [n (%)] | 1,056 (3.2%) | 4 (2.8%) | 1 |
| **Metabolic comorbidities** | |  |  |
| BMI at MRI [mean (SD)] | 26.3 (4.2) | 27.8 (4.2) | **<0.001** |
| Categories [n (%)] |  |  | **<0.001** |
| Lean (BMI < 25Kg/m^2^) | 13,869 (42%) | 34 (24%) |  |
| Overweight (BMI ≥ 25 - < 30 kg/m^2^) | 13,687 (41%) | 73 (51%) |  |
| Obese (BMI ≥ 30 Kg/m^2^) | 5,712 (17%) | 35 (25%) |  |
| Systolic blood pressure (SBP, mmHg) [mean (SD)] | 137 (19) | 144 (18) | **<0.001** |
| Categories [n (%)] |  |  | **<0.001** |
| SBP ≤ 130mmHg | 12,466 (40%) | 31 (23%) |  |
| SBP > 130mmHg | 18,552 (60%) | 102 (77%) |  |
| HbA1C (mmol/mol) [mean (SD)] | 35.0 (5.2) | 37.2 (7.1) | **<0.001** |
| Categories [n (%)] |  |  | **<0.001** |
| ≤ 42mmol/mol | 29,542 (96%) | 114 (88%) |  |
| 42mmol & < 48 mmol/mol | 787 (2.5%) | 6 (4.7%) |  |
| ≥ 48mmol/mol | 562 (1.8%) | 9 (7.0%) |  |
| LDL [mean (SD)] | 3.59 (0.83) | 3.38 (0.85) | **0.007** |
| Categories [n (%)] |  |  | 0.13 |
| ≤ 4.1 mmol/L | 23,072 (74%) | 104 (80%) |  |
| > 4.1 mmol/L | 7,997 (26%) | 26 (20%) |  |
| HDL [mean (SD)] | 1.48 (0.38) | 1.29 (0.28) | **<0.001** |
| Categories [n (%)] |  |  | 0.0501 |
| ≥ 1.04mmol/L [men] & 1.3mmol/L [women] | 23,234 (82%) | 89 (75%) |  |
| < 1.04mmol/L [men] & 1.3mmol/L [women] | 5,189 (18%) | 30 (25%) |  |
| Triglycerides [mean (SD)] | 1.63 (0.95) | 1.82 (0.92) | **0.01** |
| Categories [n (%)] | |  | 0.15 |
| ≤ 1.7 mmol/L | 20,124 (65%) | 77 (59%) |  |
| > 1.7 mmol/L | 10,991 (35%) | 54 (41%) |  |
| Total cholesterol (mmol/L) [mean (SD)] | 5.74 (1.08) | 5.35 (1.08) | **<0.001** |
| Categories [n (%)] | |  | **0.001** |
| ≤ 5.2 mmol/L | 9,917 (32%) | 59 (45%) |  |
| > 5.2 mmol/L | 21,219 (68%) | 72 (55%) |  |
| Metabolic Syndrome [n (%)] | 8,719 (35%) | 57 (52%) | **<0.001** |
| **Liver biomarkers** | |  |  |
| ALT [mean (SD)] | 23 (14) | 24 (14) | 0.38 |
| Categories [n (%)] |  |  | 0.48 |
| ≤ 45 U/L | 29,995 (96%) | 125 (95%) |  |
| > 45 U/L | 1,137 (3.7%) | 6 (4.6%) |  |
| AST [mean (SD)] | 26 (10) | 27 (9) | 0.14 |
| Categories [n (%)] |  |  | **0.01** |
| ≤ 40 U/L | 29,776 (96%) | 120 (92%) |  |
| > 40 U/L | 1,234 (4.0%) | 11 (8.4%) |  |
| FIB4 [mean (SD)] | 1.29 (0.59) | 1.55 (0.53) | **<0.001** |
| Categories [n (%)] |  |  | **<0.001** |
| FIB4 < 1.3 | 17,827 (59%) | 48 (38%) |  |
| FIB4 ≥ 1.3 & < 2.67 | 11,940 (39%) | 76 (59%) |  |
| FIB4 ≥ 2.67 | 480 (1.6%) | 4 (3.1%) |  |
| cT1 (ms) [mean (SD)] | 700 (55) | 724 (58) | **<0.001** |
| Categories [n (%)] | |  | **0.01** |
| < 800ms | 31,535 (95%) | 128 (90%) |  |
| ≥ 800ms | 1,734 (5.2%) | 14 (9.9%) |  |
| PDFF (%) [mean (SD)] | 4.9 (4.7) | 5.2 (4.2) | 0.29 |
| Categories [n (%)] |  |  | **0.01** |
| < 5% | 24,276 (73%) | 91 (64%) |  |
| ≥ 5% | 8,993 (27%) | 51 (36%) |  |
| **Cardiac biomarkers** | |  |  |
| CRP (mg/mL) [mean (SD)] | 2.00 (3.46) | 2.86 (5.80) | 0.09 |
| Categories [n (%)] |  |  | 0.15 |
| ≤ 10 mg/L | 30,272 (97%) | 125 (95%) |  |
| > 10 mg/L | 790 (2.5%) | 6 (4.6%) |  |

**S6 Table:** Characteristics of participants being hospitalised due to cardiovascular events:

|  | **CVD Hospitalization** | | |
| --- | --- | --- | --- |
|  | No CVD hospitalization (N=32,343) | CVD hospitalization (N=1,267) | P Value |
| **Demographics** | |  |  |
| Age [mean (SD)] | 64 (8) | 67 (7) | **<0.001** |
| Sex (Male) [n (%)] | 15,093 (47%) | 845 (67%) | **<0.001** |
| Race (White British) [n (%)] | 29,398 (91%) | 1,171 (92%) | 0.06 |
| Smoking [n (%)] | 1,016 (3.1%) | 50 (3.9%) | 0.108 |
| **Metabolic comorbidities** | |  |  |
| BMI at MRI [mean (SD)] | 26.3 (4.2) | 27.4 (4.2) | **<0.001** |
| Categories [n (%)] |  |  | **<0.001** |
| Lean (BMI < 25Kg/m^2^) | 13,562 (42%) | 392 (31%) |  |
| Overweight (BMI ≥ 25 - < 30 kg/m^2^) | 13,278 (41%) | 561 (44%) |  |
| Obese (BMI ≥ 30 Kg/m^2^) | 5,502 (17%) | 314 (25%) |  |
| Systolic blood pressure (SBP, mmHg) [mean (SD)] | 136 (19) | 143 (19) | **<0.001** |
| Categories [n (%)] |  |  | **<0.001** |
| SBP ≤ 130mmHg | 12,207 (41%) | 335 (28%) |  |
| SBP > 130mmHg | 17,932 (59%) | 860 (72%) |  |
| HbA1C (mmol/mol) [mean (SD)] | 35.0 (5.2) | 36.5 (6.7) | **<0.001** |
| Categories [n (%)] |  |  | **<0.001** |
| ≤ 42mmol/mol | 28,758 (96%) | 1,066 (92%) |  |
| 42mmol & < 48 mmol/mol | 754 (2.5%) | 51 (4.4%) |  |
| ≥ 48mmol/mol | 533 (1.8%) | 48 (4.1%) |  |
| LDL [mean (SD)] | 3.59 (0.83) | 3.53 (0.88) | 0.052 |
| Categories [n (%)] |  |  | 0.56 |
| ≤ 4.1 mmol/L | 22,460 (74%) | 870 (74%) |  |
| > 4.1 mmol/L | 7,748 (26%) | 312 (26%) |  |
| HDL [mean (SD)] | 1.48 (0.38) | 1.36 (0.36) | **<0.001** |
| Categories [n (%)] |  |  | **<0.001** |
| ≥ 1.04mmol/L [men] & 1.3mmol/L [women] | 22,624 (82%) | 822 (77%) |  |
| < 1.04mmol/L [men] & 1.3mmol/L [women] | 5,026 (18%) | 250 (23%) |  |
| Triglycerides [mean (SD)] | 1.63 (0.95) | 1.85 (1.01) | **<0.001** |
| Categories [n (%)] | |  | **<0.001** |
| ≤ 1.7 mmol/L | 19,643 (65%) | 655 (55%) |  |
| > 1.7 mmol/L | 10,611 (35%) | 527 (45%) |  |
| Total cholesterol (mmol/L) [mean (SD)] | 5.74 (1.08) | 5.60 (1.16) | **<0.001** |
| Categories [n (%)] | |  | **0.001** |
| ≤ 5.2 mmol/L | 9,647 (32%) | 428 (36%) |  |
| > 5.2 mmol/L | 20,627 (68%) | 756 (64%) |  |
| Metabolic Syndrome [n (%)] | 8,406 (34%) | 470 (49%) | **<0.001** |
| **Liver biomarkers** | |  |  |
| ALT [mean (SD)] | 23 (14) | 25 (14) | **<0.001** |
| Categories [n (%)] |  |  | **0.001** |
| ≤ 45 U/L | 29,175 (96%) | 1,120 (95%) |  |
| > 45 U/L | 1,095 (3.6%) | 64 (5.4%) |  |
| AST [mean (SD)] | 26 (10) | 27 (10) | **<0.001** |
| Categories [n (%)] |  |  | **<0.001** |
| ≤ 40 U/L | 28,969 (96%) | 1,107 (94%) |  |
| > 40 U/L | 1,179 (3.9%) | 74 (6.3%) |  |
| FIB4 [mean (SD)] | 1.29 (0.59) | 1.41 (0.52) | **<0.001** |
| Categories [n (%)] |  |  | **<0.001** |
| FIB4 < 1.3 | 17,408 (59%) | 551 (48%) |  |
| FIB4 ≥ 1.3 & < 2.67 | 11,531 (39%) | 576 (50%) |  |
| FIB4 ≥ 2.67 | 464 (1.6%) | 31 (2.7%) |  |
| cT1 (ms) [mean (SD)] | 700 (55) | 718 (58) | **<0.001** |
| Categories [n (%)] | |  | **<0.001** |
| < 800ms | 30,689 (95%) | 1,145 (90%) |  |
| ≥ 800ms | 1,654 (5.1%) | 122 (9.6%) |  |
| PDFF (%) [mean (SD)] | 4.9 (4.7) | 5.6 (5.1) | **<0.001** |
| Categories [n (%)] |  |  | **<0.001** |
| < 5% | 23,666 (73%) | 832 (66%) |  |
| ≥ 5% | 8,677 (27%) | 435 (34%) |  |
| **Cardiac biomarkers** | |  |  |
| CRP (mg/mL) [mean (SD)] | 2.00 (3.48) | 2.38 (3.30) | **<0.001** |
| Categories [n (%)] |  |  | **0.01** |
| ≤ 10 mg/L | 29,447 (97%) | 1,136 (96%) |  |
| > 10 mg/L | 758 (2.5%) | 43 (3.6%) |  |

**S7 Table:** Characteristics of participants deceased after MRI scan:

|  | **All-cause mortality** | | |
| --- | --- | --- | --- |
|  | Alive (n=33,374) | Death (n=236) | P Value |
| **Demographics** | |  |  |
| Age [mean (SD)] | 64 (8) | 69 (7) | **<0.001** |
| Sex (Male) [n (%)] | 15,788 (47%) | 150 (64%) | **<0.001** |
| Race (White British) [n (%)] | 30,363 (91%) | 206 (87%) | 0.04 |
| Smoking [n (%)] | 1,055 (3.2%) | 11 (4.7%) | 0.19 |
| **Metabolic comorbidities** | |  |  |
| BMI at MRI [mean (SD)] | 26.4 (4.2) | 27.2 (4.8) | **0.006** |
| Categories [n (%)] |  |  | **0.001** |
| Lean (BMI < 25Kg/m^2^) | 13,875 (42%) | 79 (33%) |  |
| Overweight (BMI ≥ 25 - < 30 kg/m^2^) | 13,743 (41%) | 96 (41%) |  |
| Obese (BMI ≥ 30 Kg/m^2^) | 5,755 (17%) | 61 (26%) |  |
| Systolic blood pressure (SBP, mmHg) [mean (SD)] | 137 (19) | 141 (19) | **<0.001** |
| Categories [n (%)] |  |  | **<0.001** |
| SBP ≤ 130mmHg | 12,476 (40%) | 66 (29%) |  |
| SBP > 130mmHg | 18,629 (60%) | 163 (71%) |  |
| HbA1C (mmol/mol) [mean (SD)] | 35.0 (5.2) | 36.9 (7.4) | **<0.001** |
| Categories [n (%)] |  |  | **<0.001** |
| ≤ 42mmol/mol | 29,633 (96%) | 191 (87%) |  |
| 42mmol & < 48 mmol/mol | 786 (2.5%) | 19 (8.7%) |  |
| ≥ 48mmol/mol | 572 (1.8%) | 9 (4.1%) |  |
| LDL [mean (SD)] | 3.58 (0.83) | 3.54 (0.86) | 0.41 |
| Categories [n (%)] |  |  | 0.52 |
| ≤ 4.1 mmol/L | 23,175 (74%) | 155 (72%) |  |
| > 4.1 mmol/L | 8,001 (26%) | 59 (28%) |  |
| HDL [mean (SD)] | 1.48 (0.38) | 1.43 (0.40) | 0.06 |
| Categories [n (%)] |  |  | 0.45 |
| ≥ 1.04mmol/L [men] & 1.3mmol/L [women] | 23,286 (82%) | 160 (80%) |  |
| < 1.04mmol/L [men] & 1.3mmol/L [women] | 5,235 (18%) | 41 (20%) |  |
| Triglycerides [mean (SD)] | 1.64 (0.95) | 1.78 (0.86) | **0.01** |
| Categories [n (%)] | |  | **<0.001** |
| ≤ 1.7 mmol/L | 20,184 (65%) | 114 (53%) |  |
| > 1.7 mmol/L | 11,038 (35%) | 100 (47%) |  |
| Total cholesterol (mmol/L) [mean (SD)] | 5.73 (1.08) | 5.65 (1.14) | 0.30 |
| Categories [n (%)] | |  | 0.42 |
| ≤ 5.2 mmol/L | 10,001 (32%) | 74 (35%) |  |
| > 5.2 mmol/L | 21,243 (68%) | 140 (65%) |  |
| Metabolic Syndrome [n (%)] | 8,783 (35%) | 93 (49%) | **<0.001** |
| **Liver biomarkers** | |  |  |
| ALT [mean (SD)] | 23 (14) | 24 (11) | 0.33 |
| Categories [n (%)] |  |  | 0.49 |
| ≤ 45 U/L | 30,087 (96%) | 208 (97%) |  |
| > 45 U/L | 1,153 (3.7%) | 6 (2.8%) |  |
| AST [mean (SD)] | 26 (10) | 27 (7) | **0.04** |
| Categories [n (%)] |  |  | 0.38 |
| ≤ 40 U/L | 29,874 (96%) | 202 (95%) |  |
| > 40 U/L | 1,242 (4.0%) | 11 (5.2%) |  |
| FIB4 [mean (SD)] | 1.30 (0.59) | 1.44 (0.53) | **<0.001** |
| Categories [n (%)] |  |  | **<0.001** |
| FIB4 < 1.3 | 17,868 (59%) | 91 (44%) |  |
| FIB4 ≥ 1.3 & < 2.67 | 11,997 (40%) | 110 (53%) |  |
| FIB4 ≥ 2.67 | 488 (1.6%) | 7 (3.4%) |  |
| cT1 (ms) [mean (SD)] | 700 (55) | 720 (59) | **<0.001** |
| Categories [n (%)] | |  | **<0.001** |
| < 800ms | 31,624 (95%) | 210 (89%) |  |
| ≥ 800ms | 1,750 (5.2%) | 26 (11%) |  |
| PDFF (%) [mean (SD)] | 4.9 (4.7) | 5.2 (5.0) | 0.28 |
| Categories [n (%)] |  |  | **0.03** |
| < 5% | 24,340 (73%) | 158 (67%) |  |
| ≥ 5% | 9,034 (27%) | 78 (33%) |  |
| **Cardiac biomarkers** | |  |  |
| CRP (mg/mL) [mean (SD)] | 2.01 (3.48) | 2.34 (3.84) | 0.21 |
| Categories [n (%)] |  |  | 0.80 |
| ≤ 10 mg/L | 30,376 (97%) | 207 (97%) |  |
| > 10 mg/L | 795 (2.6%) | 6 (2.8%) |  |

**S8 Table:** Multivariate Analysis Cox Proportional Hazard Ratios with 95% Confidence Intervals of cardiovascular outcomes, based on Z-scores to enable comparisons across different unit scales – whole population, for cases with liver IDPs were not associated with clinical outcomes.

| Metric | Outcome | HR | p value |
| --- | --- | --- | --- |
| cT1 | VTE | 1.08 (0.85 - 1.38) | 0.528 |
|  | Stroke | ns | ns |
|  | Myocardial Infarction | 1.09 (0.87 - 1.36) | 0.433 |
|  | Coronary artery disease | 1.1 (0.96 - 1.27) | 0.129 |
| PDFF | VTE | ns | ns |
|  | Stroke | ns | ns |
|  | Myocardial Infarction | 1.01 (0.81 - 1.26) | 0.938 |
|  | Coronary artery disease | 0.99 (0.86 - 1.14) | 0.849 |
| BMI | VTE | 1.05 (0.73 - 1.52) | 0.748 |
|  | Stroke | 1.17 (0.89 - 1.55) | 0.232 |
|  | Myocardial Infarction | 0.92 (0.68 - 1.23) | 0.516 |
|  | Coronary artery disease | 0.96 (0.8 - 1.15) | 0.645 |
| HbA1C | VTE | ns | ns |
|  | Stroke | 1.1 (0.96 - 1.27) | 0.103 |
|  | Myocardial Infarction | 1.11 (0.98 - 1.26) | 0.049 |
|  | Coronary artery disease | 1.12 (1.03 - 1.21) | 0.003 |
| Waist Circumference | VTE | 1.25 (0.85 - 1.85) | 0.199 |
|  | Stroke | 1.02 (0.74 - 1.41) | 0.891 |
|  | Myocardial Infarction | 1.38 (1.02 - 1.86) | 0.033 |
|  | Coronary artery disease | 1.21 (1 - 1.48) | 0.051 |
| Total cholesterol | VTE | ns | ns |
|  | Stroke | ns | ns |
|  | Myocardial Infarction | ns | ns |
|  | Coronary artery disease | ns | ns |
| AST | VTE | ns | ns |
|  | Stroke | 1.11 (0.91 - 1.36) | 0.076 |
|  | Myocardial Infarction | 1.03 (0.81 - 1.31) | 0.708 |
|  | Coronary artery disease | 1.05 (0.8 - 1.37) | 0.624 |
| ALT | VTE | ns | ns |
|  | Stroke | 0.95 (0.72 - 1.24) | 0.568 |
|  | Myocardial Infarction | 1.03 (0.76 - 1.4) | 0.82 |
|  | Coronary artery disease | 0.88 (0.65 - 1.19) | 0.34 |
| AST/ALT Ratio | VTE | ns | ns |
|  | Stroke | ns | ns |
|  | Myocardial Infarction | 1.06 (0.81 - 1.4) | 0.641 |
|  | Coronary artery disease | 0.83 (0.66 - 1.05) | 0.058 |
| C-Reactive Protein | VTE | ns | ns |
|  | Stroke | ns | ns |
|  | Myocardial Infarction | 1.08 (0.94 - 1.23) | 0.116 |
|  | Coronary artery disease | 1.03 (0.93 - 1.14) | 0.464 |
| FIB4 | VTE | ns | ns |
|  | Stroke | 0.91 (0.71 - 1.15) | 0.346 |
|  | Myocardial Infarction | 0.96 (0.75 - 1.22) | 0.678 |
|  | Coronary artery disease | 0.97 (0.83 - 1.14) | 0.688 |
| Systolic Blood Pressure | VTE | ns | ns |
|  | Stroke | 1.13 (0.95 - 1.35) | 0.152 |
|  | Myocardial Infarction | 1.18 (0.99 - 1.42) | 0.047 |
|  | Coronary artery disease | 1.11 (0.99 - 1.24) | 0.07 |
| Age at MRI | VTE | 1.39 (1.08 - 1.77) | 0.018 |
|  | Stroke | 1.96 (1.56 - 2.48) | < 0.001 |
|  | Myocardial Infarction | 1.57 (1.25 - 1.98) | < 0.001 |
|  | Coronary artery disease | 1.52 (1.32 - 1.74) | < 0.001 |
| Sex (Male) | VTE | 1.49 (0.82 - 2.68) | 0.188 |
|  | Stroke | 1.54 (0.97 - 2.44) | 0.094 |
|  | Myocardial Infarction | 1.98 (1.22 - 3.22) | 0.009 |
|  | Coronary artery disease | 1.97 (1.46 - 2.65) | < 0.001 |

Abbreviations: BMI, body mass index; HbA1C, glycated hemoglobin; ALT, alanine transaminase; AST, aspartate transaminase; LDL, low-density lipoprotein; HDL, high-density lipoprotein; CRP, c-reactive protein; cT1, iron-corrected T1 relaxation time; PDFF, proton density fat fraction; VTE, Embolism/vascular event; n.s., variables not significant in the univariate analysis were not included in the multivariate analysis.

**S9 Table: Multivariate analysis Cox proportional hazard ratios with 95% confidence intervals of cardiovascular outcomes in the subset of population with no metabolic syndrome, based on Z-scores to enable comparisons across different unit scales.**

| Metric | Outcome | HR | p value |
| --- | --- | --- | --- |
| cT1 | CVD Hospitalisation | 1.41 (1.25 - 1.58) | < 0.001 |
|  | Atrial Fibrillation | 1.2 (0.98 - 1.47) | 0.09 |
|  | Heart Failure | 1.24 (0.91 - 1.7) | 0.147 |
|  | All-Cause Mortality | ns | ns |
|  | Any CVD event | 1.04 (0.9 - 1.19) | 0.599 |
|  | VTE | ns | ns |
|  | Stroke | ns | ns |
|  | Myocardial Infarction | 1.14 (0.85 - 1.55) | 0.329 |
|  | Coronary artery disease | 1.06 (0.88 - 1.28) | 0.511 |
| PDFF | CVD Hospitalisation | 0.71 (0.61 - 0.84) | < 0.001 |
|  | Atrial Fibrillation | ns | ns |
|  | Heart Failure | ns | ns |
|  | All-Cause Mortality | ns | ns |
|  | Any CVD event | ns | ns |
|  | VTE | ns | ns |
|  | Stroke | ns | ns |
|  | Myocardial Infarction | ns | ns |
|  | Coronary artery disease | ns | ns |
| BMI | CVD Hospitalisation | 1.22 (1.04 - 1.43) | 0.012 |
|  | Atrial Fibrillation | 1.26 (0.94 - 1.7) | 0.131 |
|  | Heart Failure | ns | ns |
|  | All-Cause Mortality | ns | ns |
|  | Any CVD event | 1.06 (0.86 - 1.3) | 0.608 |
|  | VTE | 1.32 (0.76 - 2.29) | 0.361 |
|  | Stroke | ns | ns |
|  | Myocardial Infarction | 1.37 (0.87 - 2.16) | 0.153 |
|  | Coronary artery disease | ns | ns |
| HbA1C | CVD Hospitalisation | 1.12 (1.02 - 1.22) | 0.021 |
|  | Atrial Fibrillation | ns | ns |
|  | Heart Failure | 1.21 (0.97 - 1.51) | 0.128 |
|  | All-Cause Mortality | ns | ns |
|  | Any CVD event | 1.12 (1 - 1.26) | 0.094 |
|  | VTE | ns | ns |
|  | Stroke | 1.19 (0.95 - 1.49) | 0.113 |
|  | Myocardial Infarction | 1.21 (0.97 - 1.51) | 0.1 |
|  | Coronary artery disease | 1.23 (1.07 - 1.41) | 0.008 |
| Waist Circumference | CVD Hospitalisation | 1.17 (0.97 - 1.42) | 0.085 |
|  | Atrial Fibrillation | 0.95 (0.66 - 1.38) | 0.781 |
|  | Heart Failure | 1.37 (0.86 - 2.18) | 0.239 |
|  | All-Cause Mortality | ns | ns |
|  | Any CVD event | 1.19 (0.93 - 1.52) | 0.142 |
|  | VTE | 1.4 (0.69 - 2.84) | 0.32 |
|  | Stroke | 1.33 (0.89 - 1.98) | 0.078 |
|  | Myocardial Infarction | 1.12 (0.64 - 1.96) | 0.697 |
|  | Coronary artery disease | 1.15 (0.88 - 1.5) | 0.328 |
| Total Cholesterol | CVD Hospitalisation | ns | ns |
|  | Atrial Fibrillation | ns | ns |
|  | Heart Failure | ns | ns |
|  | All-Cause Mortality | ns | ns |
|  | Any CVD event | ns | ns |
|  | VTE | ns | ns |
|  | Stroke | ns | ns |
|  | Myocardial Infarction | ns | ns |
|  | Coronary artery disease | 1.19 (1.01 - 1.4) | 0.034 |
| AST | CVD Hospitalisation | 1.02 (0.88 - 1.18) | 0.673 |
|  | Atrial Fibrillation | 0.98 (0.8 - 1.18) | 0.705 |
|  | Heart Failure | 0.92 (0.62 - 1.36) | 0.666 |
|  | All-Cause Mortality | ns | ns |
|  | Any CVD event | 1.1 (0.9 - 1.33) | 0.161 |
|  | VTE | ns | ns |
|  | Stroke | 1.15 (0.95 - 1.41) | 0.048 |
|  | Myocardial Infarction | 1.08 (0.84 - 1.4) | 0.3 |
|  | Coronary artery disease | 1.15 (0.87 - 1.53) | 0.133 |
| ALT | CVD Hospitalisation | 0.99 (0.85 - 1.15) | 0.831 |
|  | Atrial Fibrillation | ns | ns |
|  | Heart Failure | ns | ns |
|  | All-Cause Mortality | ns | ns |
|  | Any CVD event | 0.84 (0.61 - 1.16) | 0.226 |
|  | VTE | ns | ns |
|  | Stroke | 1.01 (0.82 - 1.24) | 0.841 |
|  | Myocardial Infarction | 1.04 (0.8 - 1.36) | 0.598 |
|  | Coronary artery disease | 0.69 (0.42 - 1.14) | 0.118 |
| AST/ALT Ratio | CVD Hospitalisation | ns | ns |
|  | Atrial Fibrillation | ns | ns |
|  | Heart Failure | ns | ns |
|  | All-Cause Mortality | ns | ns |
|  | Any CVD event | 0.87 (0.7 - 1.08) | 0.186 |
|  | VTE | ns | ns |
|  | Stroke | ns | ns |
|  | Myocardial Infarction | ns | ns |
|  | Coronary artery disease | 0.68 (0.48 - 0.96) | 0.014 |
| C-Reactive Protein | CVD Hospitalisation | ns | ns |
|  | Atrial Fibrillation | ns | ns |
|  | Heart Failure | 1.06 (0.87 - 1.3) | 0.303 |
|  | All-Cause Mortality | ns | ns |
|  | Any CVD event | ns | ns |
|  | VTE | ns | ns |
|  | Stroke | ns | ns |
|  | Myocardial Infarction | ns | ns |
|  | Coronary artery disease | ns | ns |
| FIB4 | CVD Hospitalisation | 1 (0.9 - 1.11) | 0.973 |
|  | Atrial Fibrillation | 1.05 (0.97 - 1.14) | 0.009 |
|  | Heart Failure | 1.06 (0.91 - 1.24) | 0.046 |
|  | All-Cause Mortality | 0.96 (0.75 - 1.22) | 0.705 |
|  | Any CVD event | 1.02 (0.92 - 1.14) | 0.455 |
|  | VTE | ns | ns |
|  | Stroke | 0.92 (0.65 - 1.29) | 0.59 |
|  | Myocardial Infarction | 0.95 (0.68 - 1.34) | 0.754 |
|  | Coronary artery disease | 1.02 (0.86 - 1.21) | 0.788 |
| Systolic Blood Pressure | CVD Hospitalisation | 1.08 (0.99 - 1.19) | 0.081 |
|  | Atrial Fibrillation | 1.1 (0.93 - 1.31) | 0.241 |
|  | Heart Failure | 1.18 (0.9 - 1.55) | 0.219 |
|  | All-Cause Mortality | ns | ns |
|  | Any CVD event | 1.12 (0.99 - 1.26) | 0.057 |
|  | VTE | ns | ns |
|  | Stroke | 1.13 (0.88 - 1.45) | 0.358 |
|  | Myocardial Infarction | 1.32 (1.03 - 1.69) | 0.024 |
|  | Coronary artery disease | 1.22 (1.05 - 1.43) | 0.007 |
| Age at MRI | CVD Hospitalisation | 1.68 (1.5 - 1.88) | < 0.001 |
|  | Atrial Fibrillation | 2.2 (1.77 - 2.72) | < 0.001 |
|  | Heart Failure | 2.48 (1.7 - 3.6) | < 0.001 |
|  | All-Cause Mortality | 1.98 (1.53 - 2.55) | < 0.001 |
|  | Any CVD event | 1.74 (1.51 - 2) | < 0.001 |
|  | VTE | ns | ns |
|  | Stroke | 2.1 (1.5 - 2.95) | < 0.001 |
|  | Myocardial Infarction | 2.02 (1.41 - 2.89) | < 0.001 |
|  | Coronary artery disease | 1.59 (1.31 - 1.93) | < 0.001 |
| Sex (Male) | CVD Hospitalisation | 1.59 (1.22 - 2.07) | < 0.001 |
|  | Atrial Fibrillation | 1.85 (1.12 - 3.07) | 0.017 |
|  | Heart Failure | 3.24 (1.35 - 7.77) | 0.005 |
|  | All-Cause Mortality | ns | ns |
|  | Any CVD event | 1.6 (1.15 - 2.23) | 0.005 |
|  | VTE | 1.56 (0.57 - 4.24) | 0.388 |
|  | Stroke | 1.19 (0.61 - 2.31) | 0.592 |
|  | Myocardial Infarction | 3.15 (1.36 - 7.3) | 0.005 |
|  | Coronary artery disease | 2.15 (1.39 - 3.32) | < 0.001 |

Abbreviations: BMI, body mass index; HbA1C, glycated hemoglobin; ALT, alanine transaminase; AST, aspartate transaminase; LDL, low-density lipoprotein; HDL, high-density lipoprotein; CRP, c-reactive protein; cT1, corrected T1 relaxation time; PDFF, proton density fat fraction; Any CVD event, any cardiovascular event; VTE, Embolism/vascular event; CVD, cardiovascular; n.s., variables not significant in the univariate analysis were not included in the multivariate analysis.

**SUPLEMMENTARY REFERENCES**
